# Peripheral transcriptomic signature of chronic antibody-mediated rejection in kidney transplantation: a dual effect for *MMP9* over time

**DOI:** 10.1101/2025.11.06.25339648

**Authors:** M Morin, V Mauduit, A Bugnon, R Danger, S Brouard, A Durand, C Masset, S Ville, O Rousseau, C Kerleau, K Renaudin, G Blancho, N Vince, M Giral, S Limou

**Author notes:** Corresponding author information Pr Sophie Limou, PhD CHU Nantes Hôtel Dieu - ITUN - CR2TI 30 bd Jean Monnet F-44093 Nantes cedex 1; France.

## Abstract

**BACKGROUND:** Chronic antibody-mediated rejection (CAMR) is the main cause of late kidney allograft loss, and no specific effective treatment has been identified so far. Here, we proposed to explore the non-invasive peripheral blood transcriptome signature of CAMR.

**METHODS:** First, we compared PBMC gene expression from bulk RNA-seq between 35 patients experiencing late CAMR (mean=7.1 years) vs. 43 patients without graft dysfunction at late stages (Stable, mean=4.4 years) to identify the molecular drivers of CAMR. Second, we explored the 1-year gene expression signature in stable patients exhibiting (n=11) or not (n=51) a subsequent CAMR to define possible predictive biomarkers of CAMR.

**RESULTS:** We reported 188 differentially regulated genes during late CAMR (q<0.05). Importantly, CAMR is associated with an upregulation of genes from the degranulation pathway (*e.g. MMP9*, *MMP8* and *LCN2*) and from the C1q complement complex (*e.g. C1QA*, *C1QB* and *C1QC*), as well as with a downregulation of genes associated with subclinical rejection (*e.g. TCL1A*). The upregulated degranulation and complement signatures were validated in six independent cohorts gathering a total of 360 stable and 131 chronic rejection patients. Contrary to the injury effect observed during late stages, *MMP9* was downregulated at 1-year in PBMCs of patients who later experienced CAMR.

**CONCLUSIONS:** These results suggest a dual role for *MMP9* expression with an early protective effect against CAMR and deleterious effects in the later stage. *MMP9* peripheral expression appears as a promising biomarker candidate for kidney transplantation follow-up.

**Translational Statement:** CAMR is the main cause of late kidney allograft loss, and we aimed to identify molecular targets and biomarkers. By comparing CAMR and stable kidney-transplanted patients prior and during diagnosis, we identified *MMP9* as a potential biomarker for both CAMR prognosis and diagnosis. We also highlighted the C1q complement complex and *TCL1A* in CAMR as potential diagnostic biomarkers. These results provide new insights into CAMR pathophysiology and may guide the development of innovative treatment targeting *MMP9* expression in kidney transplanted patients. Ultimately, this work laid the foundation for exploring *MMP9* expression kinetics and develop new ways of treating CAMR patients.

## Introduction

Chronic antibody-mediated rejection (CAMR) is the primary cause of late kidney graft failure and is currently defined by three key factors according to the Banff classification^1^: histologic evidence of chronic tissue injury such as transplant glomerulopathy; evidence of ongoing or recent antibody interaction with the endothelium, indicated by C4d deposition; and serologic evidence of donor-specific antibodies (DSA)^1^. However, despite these criteria, the molecular triggers underlying CAMR remain largely elusive and require further investigation. Understanding these mechanisms is crucial for developing more effective therapeutic strategies.

Over the past decade, numerous research groups have attempted to clarify the peripheral blood mononuclear cells (PBMC) transcriptomic profile associated with kidney allograft rejection, primarily focusing on antibody-mediated rejection (AMR). AMR is an acute alloimmune injury driven by DSA that can evolve into CAMR. Van Loon *et al.* described an interferon-regulatory signature for AMR, alongside genes involved in the antigen presentation or the complement pathways^2^. Importantly, combination between biopsy and peripheral blood transcriptomics showed that the two are closely correlated in kidney-transplanted patients, highlighting the relevance of investigating the non-invasive blood compartment^2^. Two studies specifically focused on CAMR and identified an upregulation of several immune-related genes^2^ and a NFκB signaling enrichment^3^, respectively. However, these studies focused on a low number of patients (29 vs. 29 using microarrays^4^ and 2 vs. 2 using scRNA-seq^3^). PBMC transcriptomic also contributed to define a minimal peripheral AMR diagnostic signature from 8 genes^5^, and a composite diagnostic subclinical rejection score^6^. The latter integrated *AKR1C3* and *TCL1A* gene expressions alongside clinical factors (previous rejection episodes, previous transplantation(s), recipient sex and tacrolimus uptake)^6^.

In our study, we proposed to delve into the molecular networks underlying CAMR from PBMC RNA-seq data to address the knowledge gap of this major cause of graft loss.

## Methods

### Sample selection

Samples were extracted from the DIVAT multicentric biocollection^7^ (approved by the CNIL [DR-2025-087 N°914184, February 15, 2015; ClinicalTrials.gov recording NCT02900040] and the French Ministry of Higher Education and Research [file 13.334-cohort DIVAT RC12_0452, www.divat.fr]), which gathers clinical data and biological samples (*e.g.* blood, biopsies, urine) for kidney transplanted patients. We selected 140 Trizol-stored PBMC samples from the Nantes center, representing 35 CAMR samples, 43 Stable samples and 62 Reference samples (Figure 1, see Supplementary Methods).

**Figure 1:**
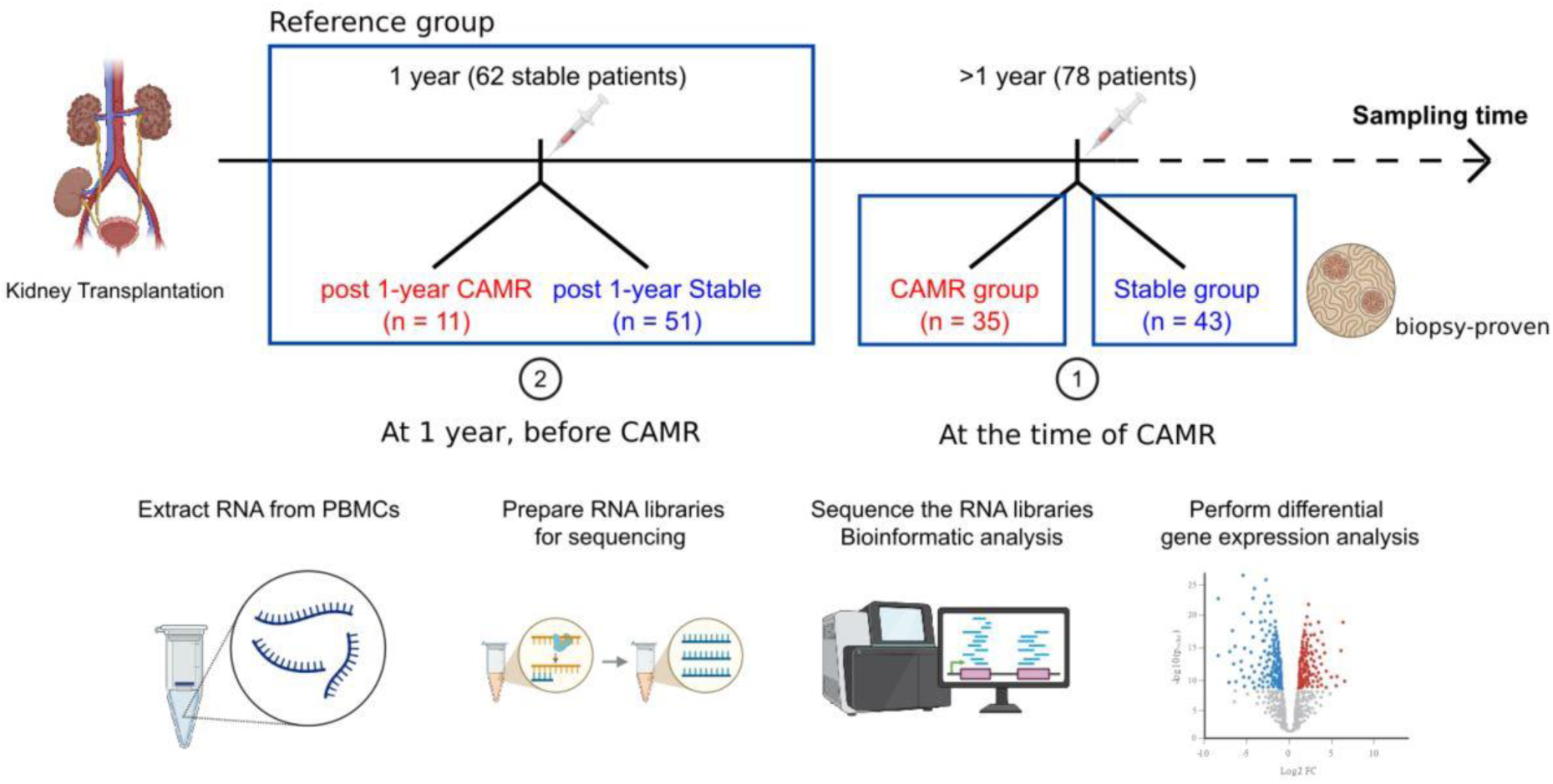
Study design. We selected 140 PBMC samples from kidney-transplanted patients for whom we performed RNA-seq. First, we selected 78 PBMCs sampled at the time of biopsy evaluation more than 1 year after transplantation, and divided into 35 CAMR patients (defined according to Banff2022) and 43 Stable patients (with stable kidney function under treatment and no current or previously recorded rejection event). Comparing these two groups defined a peripheral CAMR diagnosis signature. Second, we sampled 62 Reference PBMCs from patients with stable kidney function at 1-year post-transplantation and no biopsy-proven rejection during the first-year post-transplantation. Within this group, 11 subsequently experienced a CAMR event and 51 remained stable. Patients with less than 12.5 years of follow-up were censured in time-dependant analysis. Comparing these two Reference subgroups defined an early subclinical CAMR signature. For all PBMC samples, we isolated RNA, prepared and sequenced the RNA libraries, and ran the bioinformatic analyses to identify differentially expressed genes (see Methods and Supplementary Methods).

### Total RNA-seq preparation and analysis

Library preparations and total RNA-seq were outsourced to Parean biotechnologies (Saint-Malo, France). Our analytic pipeline was implemented using Snakemake (version 7.14.1) and is available at the following repository: https://gitlab.univ-nantes.fr/morin-m-2/imokit. Two differential expression analyses were performed (Figure 1): 1) comparing CAMR (n=35) vs. Stable (n=43) patients, and 2) comparing Reference patients who experienced a CAMR after 1 year (n=11) vs. Reference patients who did not (n=51). All transcriptomic analyses were corrected for patients’ sex, age and BMI at sampling, post-transplantation time at sampling, sequencing line, and graft year. Differentially expressed genes were defined by a q-value (false discovery rate, FDR) below 0.05. For further technical details, see the Supplementary Methods.

### Deconvolution and synthetic scRNA-seq

Single-cell RNA-seq (scRNA-seq) data (n=15,263 cells) were obtained from the Gene Expression Omnibus (GEO) database under accession number GSE230651^8^ for 1-year PBMC samples of kidney-transplanted patients categorized as tolerant (n=1) or stable (n=2). Cell deconvolution was conducted from these reference scRNA-seq data using the online CIBERSORTx server^9^ to impute cell proportions and perform a synthetic scRNA-seq analysis of our bulk data (see Supplementary Methods).

### Cox regression models and time-dependent analyses

From the 62 Reference samples, 11 patients later experienced a CAMR episode during the first 12.5 years post-transplantation (mean=5.8 years). The Cox proportional hazards multivariable model was adjusted for graft year, recipient’s and donor’s age, sex, BMI, as well as for 1-year kidney graft function computed with the CKD-EPI equation, *HLA* ABDR incompatibility, induction treatment, CNI dose, dialysis duration, and donor’s creatinine levels. These covariates were selected based on their clinical relevance to CAMR diagnosis (*e.g.* kidney graft function, *HLA* ABDR incompatibility) or for their association with specific transcriptomic pattern (*e.g.* age, sex). We tested the association between the 1-year *MMP9* expression and time-to-CAMR using the survival R package (version 3.5). A Kaplan Meier curve was produced using the survminer package (version 0.5.0) by discriminating between low vs. high *MMP9* expression levels according to the median *MMP9* expression.

### Independent validation of the CAMR diagnosis signature

We explored transcriptomic data from the GEO database. We only extracted chronic rejection (CR, n=131) and stable (n=360) kidney-transplanted patients with PBMC or blood samples (total of 6 studies): GSE14655^10^, GSE222889^11^, GSE22707^12^, GSE47755^13^, GSE50084^14^, and GSE51675^4^ (Table 1).

**Table 1:** Characteristics of the six independent validation datasets used in the CAMR diagnosis meta-analysis. The first column corresponds to the accession number in the GEO portal. The sample type (PBMC or whole blood), experimental technology (Array or RNA-seq), and numbers of Stable and CR samples are displayed for each dataset. We also reported the mean post-transplantation collecting time when available. *All CR samples correspond to biopsy-proven CAMR samples, except for GSE14655 (chronic rejection events with no further details on immune cell infiltrates). *yr, years; NA, not available.*

| **GEO**<br>**accession**<br>**number** | **Sample type** | **Experiment** | **Stable** | **Time at**<br>**draw** | **Chronic**<br>**Rejection** | **Time at**<br>**draw** |
| --- | --- | --- | --- | --- | --- | --- |
| *GSE14655* | Whole blood | Array | 100 | 6 yrs | 35 | 5 yrs |
| *GSE222889* | Whole blood | RNA-seq | 23 | 24 yrs | 16* | 6 yrs |
| *GSE22707* | PBMC | Array | 12 | 8 yrs | 12* | 9 yrs |
| *GSE47755* | PBMC | Array | 190 | NA | 31* | NA |
| *GSE50084* | Whole blood | Array | 26 | <1 yr | 28* | 4 yrs |
| *GSE51675* | PBMC | Array | 9 | 3 yrs | 9* | 4 yrs |

We only retained genes with data available in ≥3 studies and patients with <33% of missing data (see Supplementary Methods).

## Results

### MMP9, TCL1A and C1Q complement complex genes were differentially expressed during CAMR

To identify CAMR transcriptomic patterns in PBMC, we compared 35 CAMR vs. 43 Stable patients (Table 2). The mean sampling time was 2.7 years later in the CAMR group (average time: 2603 days post-transplantation, *i.e.* 7.1 years) than in the Stable group (average time: 1622 days post-transplantation, *i.e.* 4.4 years). Both groups were comparable for most demographics and clinical characteristics (significant pvalues <0.001 when accounting for multiple testing). Recipients and donors were mostly male (66% and 60%, respectively) and were nominally younger in the CAMR than Stable group (42 vs. 49 and 44 vs. 53, respectively). In our study, 78% of our patients were transplanted for the first time, and there was an average of 3 out 6 *HLA* ABDR mismatches between donors and recipients. Treatments were similar between the two groups with a nominally higher CNI dose and higher proportion of corticosteroid treatment for the CAMR patients. As expected, CAMR patients significantly presented more often with *de novo* DSA (73% vs. 29%), with higher g, ptc and cg lesions (P<0.001), and with more C4d staining (67% vs. 3%) than Stable patients, in accordance with the Banff CAMR definition.

**Table 2:**
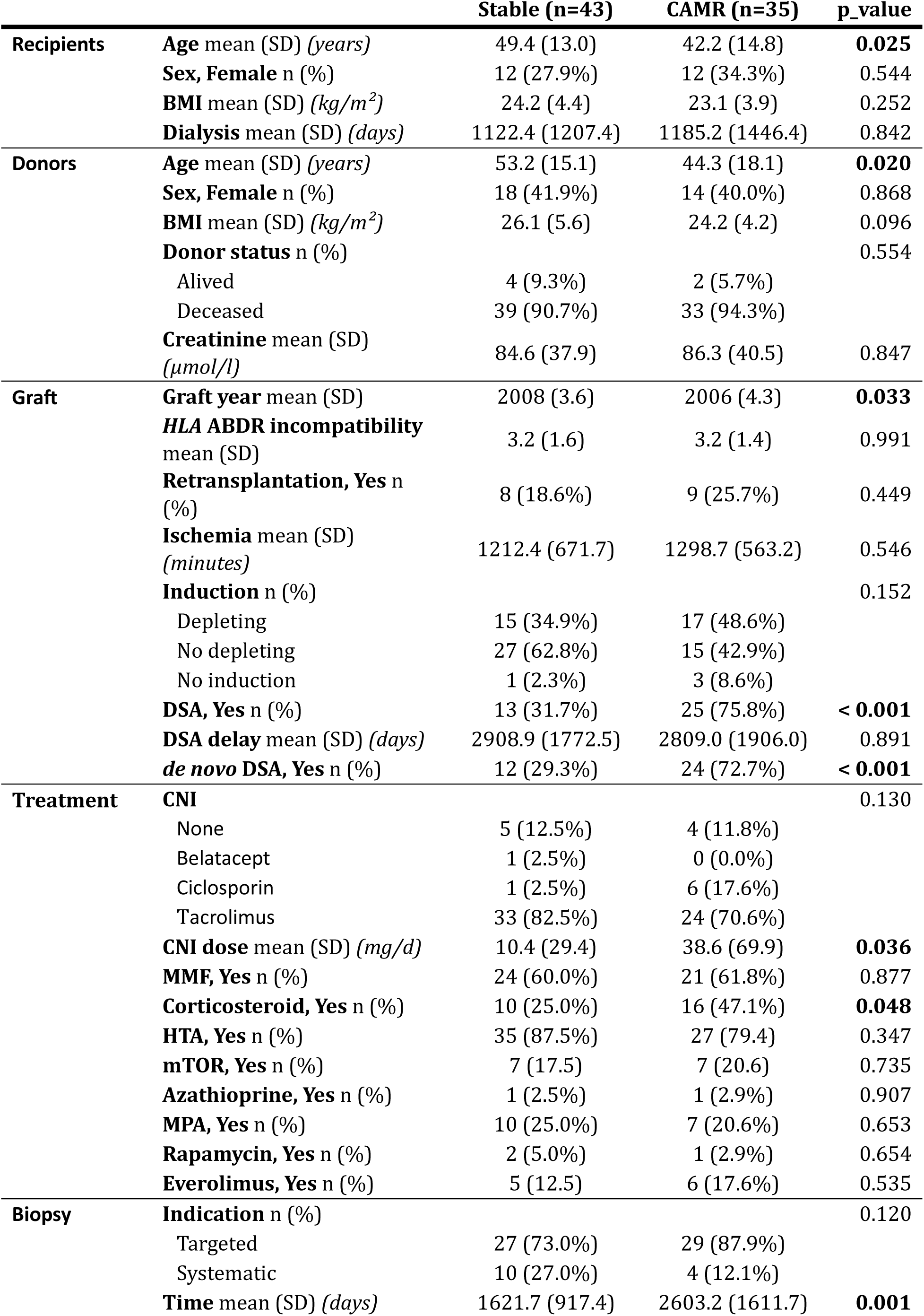
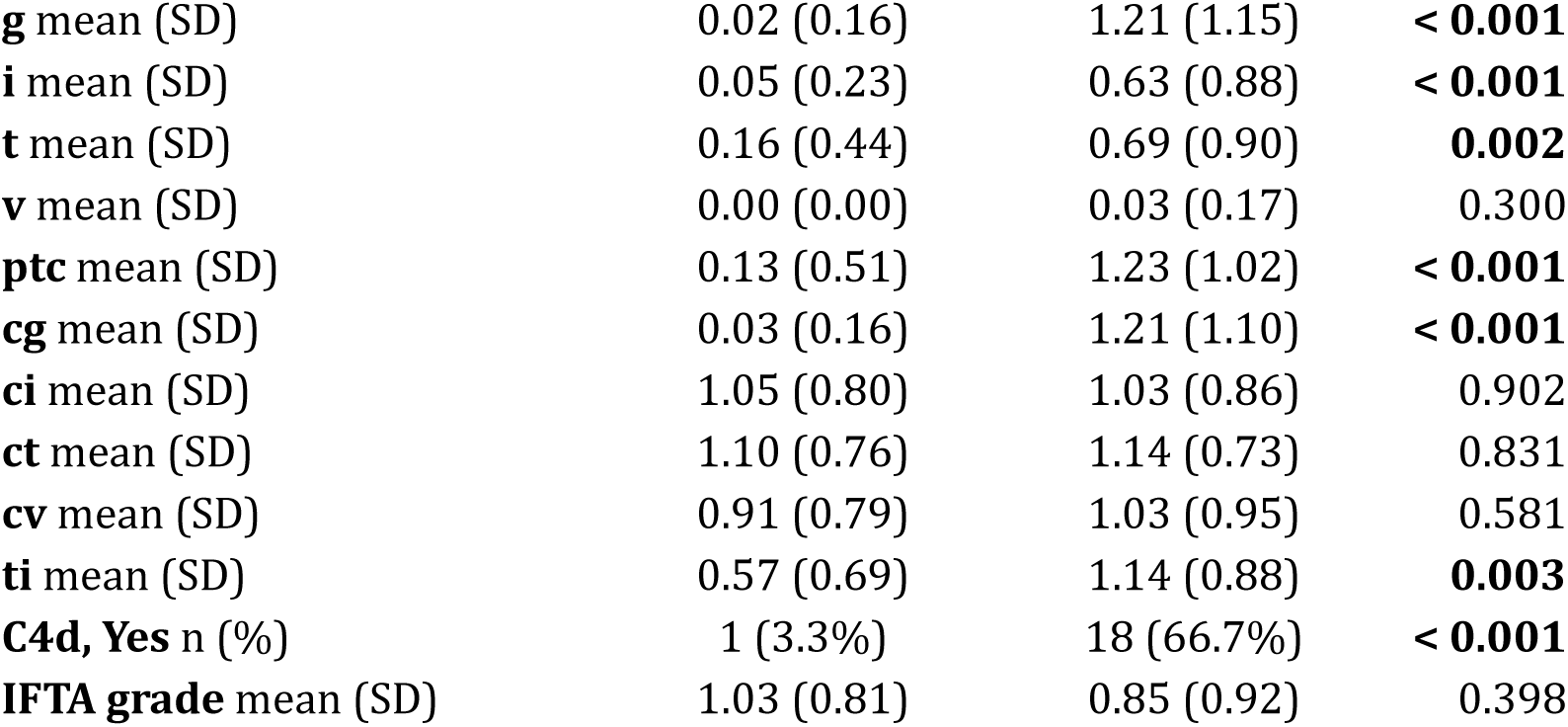
Characteristics of the late Stable and CAMR patients. Categorical data (*e.g.* sex, retransplantation) are presented in absolute values and percentages, and were compared between both groups of interest with a chi-squared test. Continuous data (*e.g.* age, BMI) are presented as mean with standard deviation (SD), and were tested between both groups using an ANOVA t-test with equal variances. Significance was defined for pvalues <0.001 to account for multiple testing. Nominally significant P-values<0.05 are highlighted in bold. *BMI, Body mass index; DSA, donor-specific antibodies; CNI, Calcineurin inhibitor; MMF, Mycophenolate mofetil; HTA, high blood pressure treatment; MPA, Mycophenolic Acid; g, acute glomeruli lesions; i, acute interstitial lesions; t, acute tubular lesions; v, acute vascular lesions; ptc, peritubular capillaritis; cg, chronic glomeruli lesions; ci, chronic interstitial lesions; ct, chronic tubular lesions; cv, chronic vascular lesions; ti, total inflammation score; IFTA, interstitial fibrosis and tubular atrophy.*

|  |  | Stable (n=43) | CAMR (n=35) | p_value |
| --- | --- | --- | --- | --- |
| **Recipients** | **Age** mean (SD) (*years*) | 49.4 (13.0) | 42.2 (14.8) | **0.025** |
|  | **Sex, Female** n (%) | 12 (27.9%) | 12 (34.3%) | 0.544 |
|  | **BMI** mean (SD) (*kg/m<sup>2</sup>*) | 24.2 (4.4) | 23.1 (3.9) | 0.252 |
|  | **Dialysis** mean (SD) (*days*) | 1122.4 (1207.4) | 1185.2 (1446.4) | 0.842 |
| **Donors** | **Age** mean (SD) (*years*) | 53.2 (15.1) | 44.3 (18.1) | **0.020** |
|  | **Sex, Female** n (%) | 18 (41.9%) | 14 (40.0%) | 0.868 |
|  | **BMI** mean (SD) (*kg/m<sup>2</sup>*) | 26.1 (5.6) | 24.2 (4.2) | 0.096 |
|  | **Donor status** n (%) |  |  | 0.554 |
|  | Alived | 4 (9.3%) | 2 (5.7%) |  |
|  | Deceased | 39 (90.7%) | 33 (94.3%) |  |
|  | **Creatinine** mean (SD) (*μmol/l*) | 84.6 (37.9) | 86.3 (40.5) | 0.847 |
| **Graft** | **Graft year** mean (SD) | 2008 (3.6) | 2006 (4.3) | **0.033** |
|  | **HLA ABDR incompatibility** mean (SD) | 3.2 (1.6) | 3.2 (1.4) | 0.991 |
|  | **Retransplantation, Yes** n (%) | 8 (18.6%) | 9 (25.7%) | 0.449 |
|  | **Ischemia** mean (SD) (*minutes*) | 1212.4 (671.7) | 1298.7 (563.2) | 0.546 |
|  | **Induction** n (%) |  |  | 0.152 |
|  | Depleting | 15 (34.9%) | 17 (48.6%) |  |
|  | No depleting | 27 (62.8%) | 15 (42.9%) |  |
|  | No induction | 1 (2.3%) | 3 (8.6%) |  |
|  | **DSA, Yes** n (%) | 13 (31.7%) | 25 (75.8%) | **< 0.001** |
|  | **DSA delay** mean (SD) (*days*) | 2908.9 (1772.5) | 2809.0 (1906.0) | 0.891 |
|  | **de novo DSA, Yes** n (%) | 12 (29.3%) | 24 (72.7%) | **< 0.001** |
| **Treatment** | **CNI** |  |  | 0.130 |
|  | None | 5 (12.5%) | 4 (11.8%) |  |
|  | Belatacept | 1 (2.5%) | 0 (0.0%) |  |
|  | Ciclosporin | 1 (2.5%) | 6 (17.6%) |  |
|  | Tacrolimus | 33 (82.5%) | 24 (70.6%) |  |
|  | **CNI dose** mean (SD) (*mg/d*) | 10.4 (29.4) | 38.6 (69.9) | **0.036** |
|  | **MMF, Yes** n (%) | 24 (60.0%) | 21 (61.8%) | 0.877 |
|  | **Corticosteroid, Yes** n (%) | 10 (25.0%) | 16 (47.1%) | **0.048** |
|  | **HTA, Yes** n (%) | 35 (87.5%) | 27 (79.4) | 0.347 |
|  | **mTOR, Yes** n (%) | 7 (17.5) | 7 (20.6) | 0.735 |
|  | **Azathioprine, Yes** n (%) | 1 (2.5%) | 1 (2.9%) | 0.907 |
|  | **MPA, Yes** n (%) | 10 (25.0%) | 7 (20.6%) | 0.653 |
|  | **Rapamycin, Yes** n (%) | 2 (5.0%) | 1 (2.9%) | 0.654 |
|  | **Everolimus, Yes** n (%) | 5 (12.5) | 6 (17.6%) | 0.535 |
| **Biopsy** | **Indication** n (%) |  |  | 0.120 |
|  | Targeted | 27 (73.0%) | 29 (87.9%) |  |
|  | Systematic | 10 (27.0%) | 4 (12.1%) |  |
|  | **Time** mean (SD) (*days*) | 1621.7 (917.4) | 2603.2 (1611.7) | **0.001** |

|  |  |  |  |
| --- | --- | --- | --- |
| **g** mean (SD) | 0.02 (0.16) | 1.21 (1.15) | **< 0.001** |
| **i** mean (SD) | 0.05 (0.23) | 0.63 (0.88) | **< 0.001** |
| **t** mean (SD) | 0.16 (0.44) | 0.69 (0.90) | **0.002** |
| **v** mean (SD) | 0.00 (0.00) | 0.03 (0.17) | 0.300 |
| **ptc** mean (SD) | 0.13 (0.51) | 1.23 (1.02) | **< 0.001** |
| **cg** mean (SD) | 0.03 (0.16) | 1.21 (1.10) | **< 0.001** |
| **ci** mean (SD) | 1.05 (0.80) | 1.03 (0.86) | 0.902 |
| **ct** mean (SD) | 1.10 (0.76) | 1.14 (0.73) | 0.831 |
| **cv** mean (SD) | 0.91 (0.79) | 1.03 (0.95) | 0.581 |
| **ti** mean (SD) | 0.57 (0.69) | 1.14 (0.88) | **0.003** |
| **C4d, Yes** n (%) | 1 (3.3%) | 18 (66.7%) | **< 0.001** |
| **IFTA grade** mean (SD) | 1.03 (0.81) | 0.85 (0.92) | 0.398 |

The differential expression analysis identified 67 downregulated and 121 upregulated genes in CAMR compared to Stable patients (Figure 2A, Suppl. Table 1). The unsupervised clustering of patients from these 188 differentially expressed genes (DEG) demonstrated a limited separation between CAMR and Stable samples (Figure 2B). The pathway enrichment analysis revealed a significant enrichment for multiple immune system pathways and extracellular matrix remodelling pathways in the significant DEG (Figure 2C, Suppl. Table 2), including the degranulation (p=2.3×10^-21^), C1q complement complex (p=2.5×10^-7^) and chemokine signaling (p=3.4×10^-6^) pathways.

**Figure 2:**
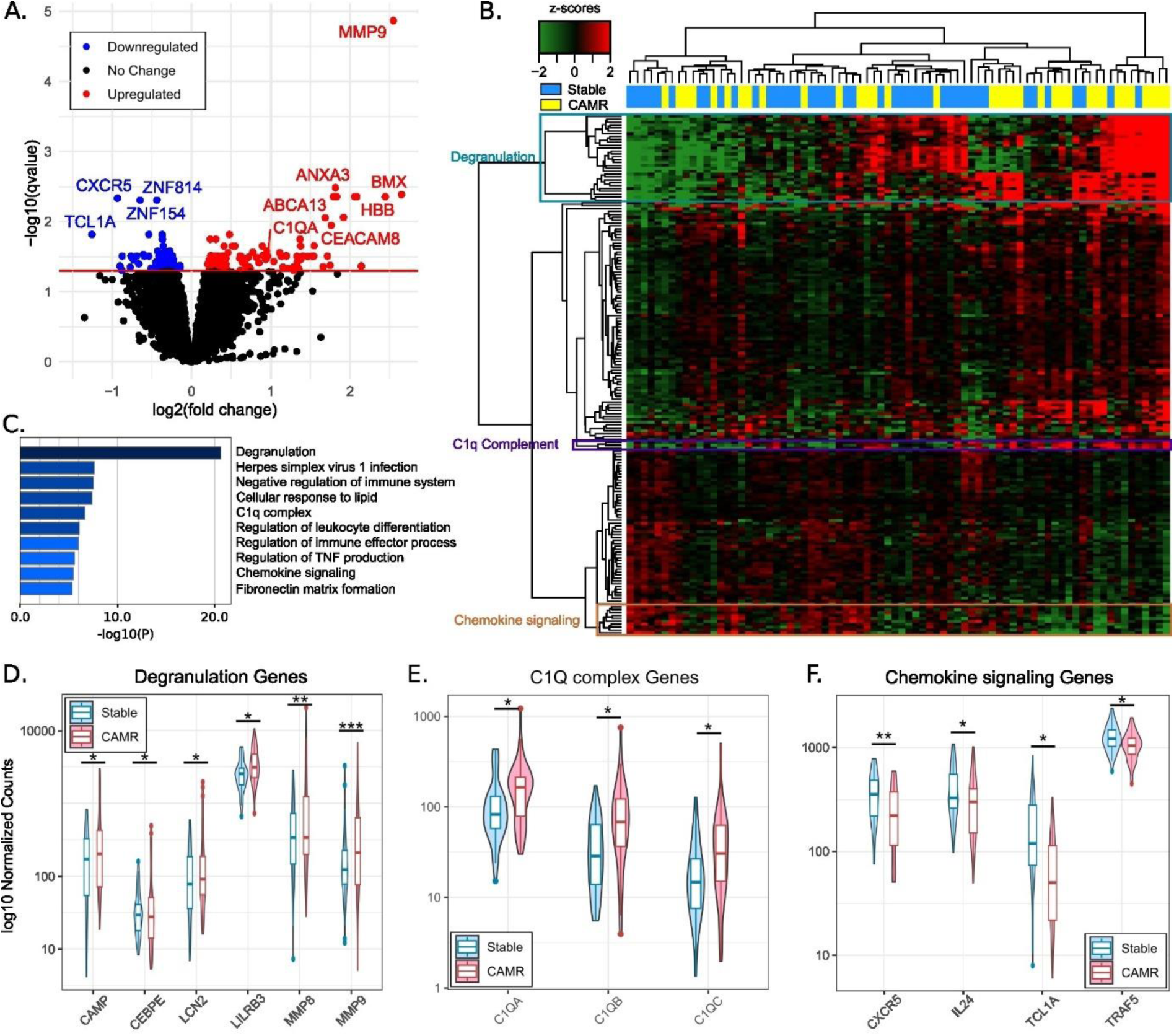
Gene expression changes in PBMCs of Stable vs. CAMR late patients. **A.** The volcano plot shows the log_2_(Fold Change) and the -log_10_(qvalue) of the 14,203 genes tested for differential expression. Genes colored in blue (67) and red (121) were significantly downregulated and upregulated, respectively (qvalue≤0.05). **B.** The heatmap displays the expression levels in z-score for significant genes within each sample. Red indicates upregulation, while green indicates downregulation within the CAMR samples. Sample and gene clustering are shown on the top and on the left side respectively. Genes associated with the degranulation pathway are highlighted in turquoise, with the C1q complement in purple, and with the chemokine signaling in ochre. **C.** Pathway enrichment analysis of significant genes. The bar plot is colored according to the p-value (dark blue: -log_10_(p-value)≥20, blue: -log_10_(p-value)≥6, and light blue: -log_10_(p-value)≥2). Violin plots display the normalized expression of genes associated with the degranulation (**D**), C1q complement (**E**) and chemokine signaling (**F**) pathways. *qvalue≤0.05, **qvalue≤0.01, and ***qvalue≤0.001.

Focusing on the degranulation/extracellular matrix top pathway, we systematically reported a higher gene expression within the CAMR samples (Figure 2D), including *MMP9* (log_2_(FC)=2.55, q-value=1.36×10^-5^), *MMP8* (log^2^(FC)= 2.06, q-value=4.41×10^-3^) and *LCN2* (log_2_(FC)= 1.41, q-value= 3.11×10^-2^). *CAMP* (log_2_(FC)=1.55, q-value=2.20×10^-2^), encoding an enzyme associated with the MMP9 and MMP8 metalloproteinases, *LILRB3* (log_2_(FC)=0.61, q-value=3.30×10^-2^), previously associated with kidney transplant failure, and *CEBPE* (log_2_(FC)=1.13, q-value=3.10×10^-2^), encoding a transcription factor regulating *MMP9* expression, were also upregulated in CAMR samples. Importantly, the three complement genes involved in the C1q complex that initiates the classical complement cascade and C4 cleavage, namely *C1QA* (log_2_(FC)=0.97, q-value=3.11×10^-2^), *C1QB* (log_2_(FC)=1.37, q-value=1.78×10^-2^) and *C1QC* (log_2_(FC)=1.38, q-value=3.45×10^-2^), were all upregulated in CAMR samples (Figure 2E).

Finally, we observed a systematic decreased expression of genes encoding negative regulators of immune responses and involved in chemokine signaling in CAMR, such as *IL24* (log_2_(FC)=-0.76, q-value=3.11×10^-2^), *CXCR5* (log_2_(FC)=-0.93, q-value=4.63×10^-3^) and *TRAF5* (log_2_(FC)=-0.37, q-value=1.52×10^-2^) (Figure 2F). Interestingly, *TCL1A*, which expression was previously reported as lower during subclinical rejection^6^, was also downregulated in CAMR (log_2_(FC)=-1.26, q-value=1.52×10^-2^).

### Cell-specific gene expression signature of CAMR

PBMCs are composed of a variety of cell types, each expressing their own specific transcriptome. We therefore evaluated if cellular unbalance could have influenced our bulk RNA-seq-based signature of CAMR using PBMC scRNA-seq data (see Methods). First, we annotated the cell clusters (Suppl. Figure 1) and observed, in accordance with previous reports^8^, that *TCL1A* was predominantly expressed in naive B cells and plasmacytoid dendritic cells, while the C1q genes were primarily expressed in CD16^+^ non-classical monocytes (Figure 3A). *MMP9* and associated genes were mainly expressed in CD14^+^ classical monocytes of kidney recipients (Figure 3B), with nearly no detectable expression in healthy individuals (Suppl. Figure 2).

**Figure 3:**
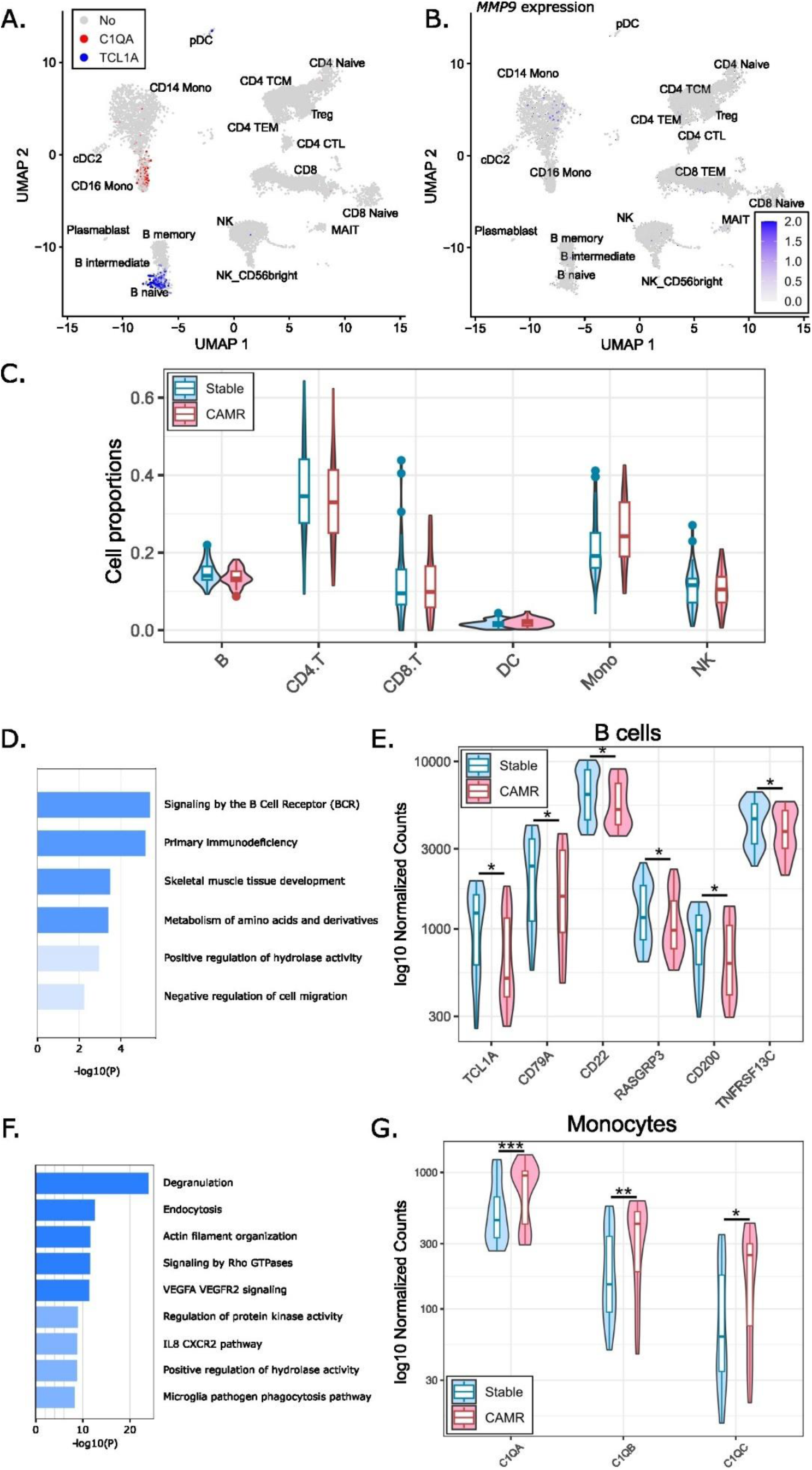
Cell-specific expression of our top CAMR genes. **A.** Cell-specific expression of the *TCL1A* (blue) and *C1QA* (red) genes within kidney recipients’ scRNAseq data. **B.** Cell-specific expression intensity (shades of blues) of *MMP9* within kidney recipients’ scRNAseq data. **C.** Violin plots showing the PBMC cell proportions imputed from bulk RNA-seq data for Stable (blue) and CAMR (red) patients. No cell type exhibited a significant difference between the Stable and CAMR groups. **D.** Pathway enrichment analysis results for the DEG identified in the synthetic B cell compartment. Blue and light blue bars indicate p≤0.001 and p≤0.01, respectively. **E.** Violin plots showing the normalized expression of BCR signaling-associated genes in the synthetic B cell compartment. **F.** Pathway enrichment analysis results for the DEG identified in the synthetic monocyte compartment. Dark blue, blue and light blue bars indicate p≤10^-10^, p≤0.001 and p≤0.01, respectively. **G.** Violin plots showing the normalized expression of the C1q complement complex-associated genes in the synthetic monocyte compartment. *qvalue≤0.05, **qvalue≤0.01, and ***qvalue≤0.001. *Mono, monocyte; DC, dendritic cell; pDC, Plasmacytoid dendritic cells; cDC2, classical dendritic cell 2; TCM, T central memory cell; TEM, T effector memory cell; CTL, cytotoxic T lymphocyte; MAIT, mucosal associated invariant T cell; DEG, differentially expressed genes*.

Second, we performed a cell deconvolution to assess the cell composition of our CAMR and Stable PBMC samples. The cell proportions aligned closely with what is typically expected in PBMCs and we did not observe any significant differences in cell proportions between CAMR and Stable patients (Figure 3C, Suppl. Figure 3, Suppl. Table 3 and 4).

Finally, we performed a synthetic scRNA-seq analysis of CAMR from our bulk RNA-seq data using cell supertype annotations. In the B cell synthetic compartment, we observed a downregulation of *TCL1A* (log_2_(FC)=-0.57, q-value=1.50×10^-2^), along with other genes involved in the BCR signaling pathway such as *CD79A*, *CD22*, *RASGRP3*, *CD200* and *TNFRSF13C* (Figures 3D and 3E). Conversely, within the monocyte synthetic compartment, we identified an upregulation of the C1q complement complex genes such as *C1QA* (log_2_(FC)=0.63, q-value=5.62×10^-4^), *C1QB* (log_2_(FC)=0.81, q-value=1.58×10^-3^) and *C1QC* (log_2_(FC)=0.87, q-value=1.64×10^-2^) (Figures 3F and 3G). We could not build a prediction model for some low-expression genes in our scRNA-seq dataset, such as *MMP9* or other associated genes including *MMP8* or *LCN2* (Figure 3B, Suppl. Figure 2). To overcome the limited number of *MMP9*-expressing cells in our scRNA-seq reference dataset, we explored the Tabula Sapiens public dataset^15^ and confirmed that *MMP9* is mainly expressed in a monocyte’s subgroup in PBMCs (Suppl. Figure 4).

Overall, these data strongly support that the transcriptomic signatures and conclusions drawn from the bulk RNA-seq analysis are not biased by unbalanced cell type proportions and reflect real differential gene expression profiles during CAMR.

### Rejection subtype specificity of the identified differentially expressed genes

We next wanted to explore whether the top DEG were CAMR-specific or could also be associated with other rejection subtypes. Indeed, we previously observed that the CAMR clinical hallmarks (DSA, C4d deposition, and cg, g and ptc lesions) were, as expected, significantly enriched in our CAMR patients, but also reported a significant enrichment for i lesions (p<0.001) and nominal enrichments for the t (p=0.002) and ti (p=0.003) lesions (Table 2), which are histopathologic markers used for the T-cell mediated rejection (TCMR) diagnosis.

Using the same statistical models as in the discovery Stable vs. CAMR analysis, we therefore tested the association of our top DEG expression with clinical markers for rejection diagnosis. As expected, our top genes were significantly associated with C4d staining, a marker of AMR (Figure 4A), and with cg lesions, a marker of chronicity for AMR (Figure 4B), with effect sizes in the same direction as in the initial CAMR analysis. On the other hand, only *C1QB* and *TCL1A* genes were nominally associated with t and i lesion scores (Figures 4C and 4D), suggesting that the identified DEG would in part be specific to the AMR/CAMR rejection subtypes.

**Figure 4:**
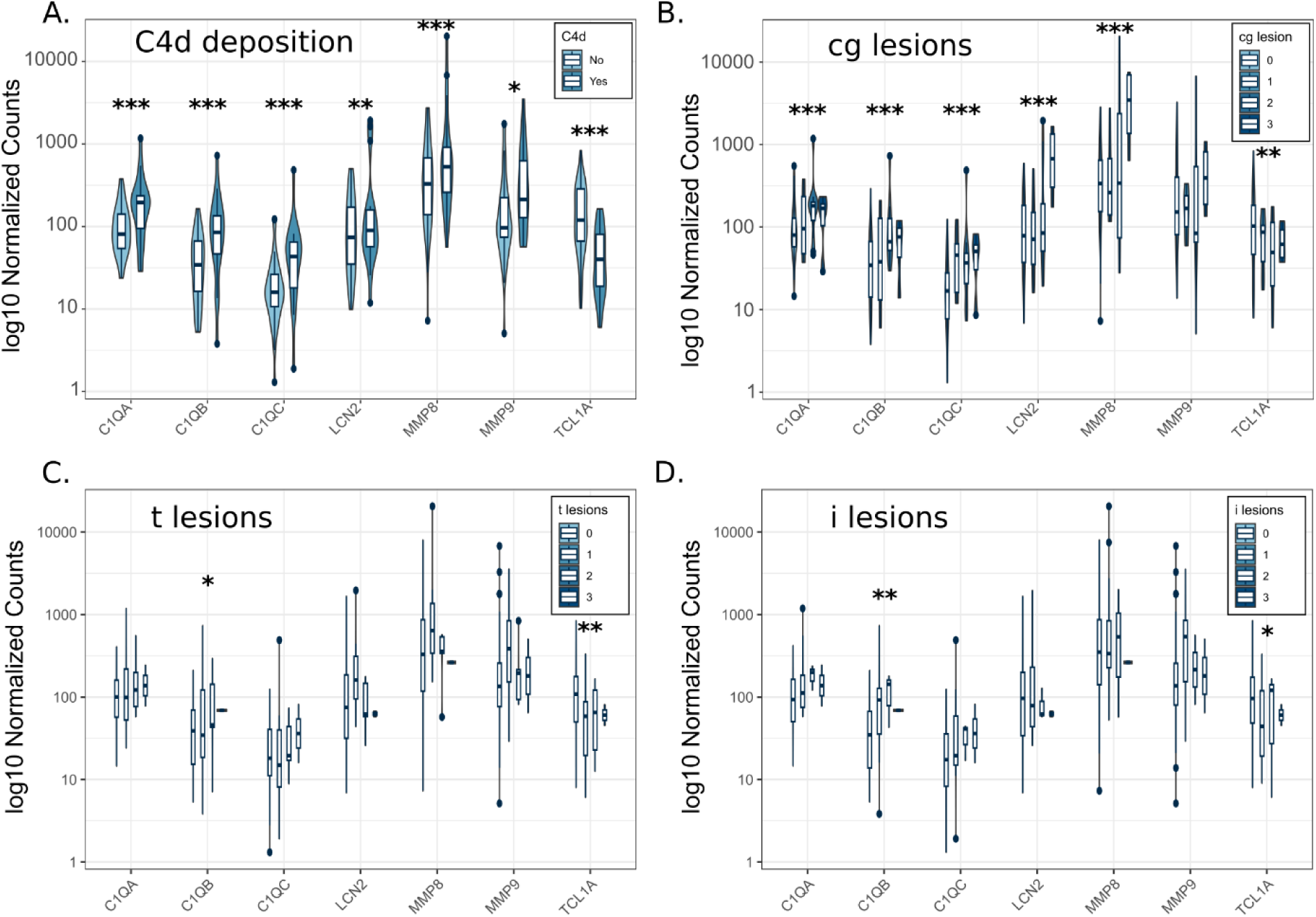
Top genes association with rejection clinical markers. Violin plots presenting the association of some top genes PBMC normalized expression (*C1QA*, *C1QB*, *C1QC*, *LCN2*, *MMP8*, *MMP9* and *TCL1A*) with C4d deposition (**A**), cg histologic lesion scores (**B**), t lesion scores (**C**), and i lesion scores (**D**). *qvalue≤0.05, **qvalue≤0.01, and ***qvalue≤0.001. *cg, chronic glomerulitis; t, tubulitis; i, interstitial inflammation*.

### Independent validation of the CAMR signature

In order to validate our results, we collected transcriptomic data from six independent studies with PBMC (n=3, total of 211 Stable vs. 52 CR) or whole blood (n=3, total of 149 Stable vs. 79 CR) samples from kidney transplanted patients (Table 1). 96 out of 131 (73%) CR samples corresponded to biopsy-proven CAMR. We performed a differential expression meta-analysis for the DEG described above (Figure 5A). Using an FDR threshold of 0.05, we validated 33 out of 176 (19%) genes, including *C1QA* (q-value=2.21×10^-2^), *C1QB* (q-value=1.55×10^-3^), *C1QC* (q-value=3.85×10^-2^) and *MMP9* (q-value=2.40×10^-2^), which were all upregulated in the meta-analysis (Figure 5B). Altogether, these 33 validated genes represent top priority candidates to further explore the CAMR pathophysiology mechanisms and their CAMR biomarker potential (Suppl Table 5).

**Figure 5:**
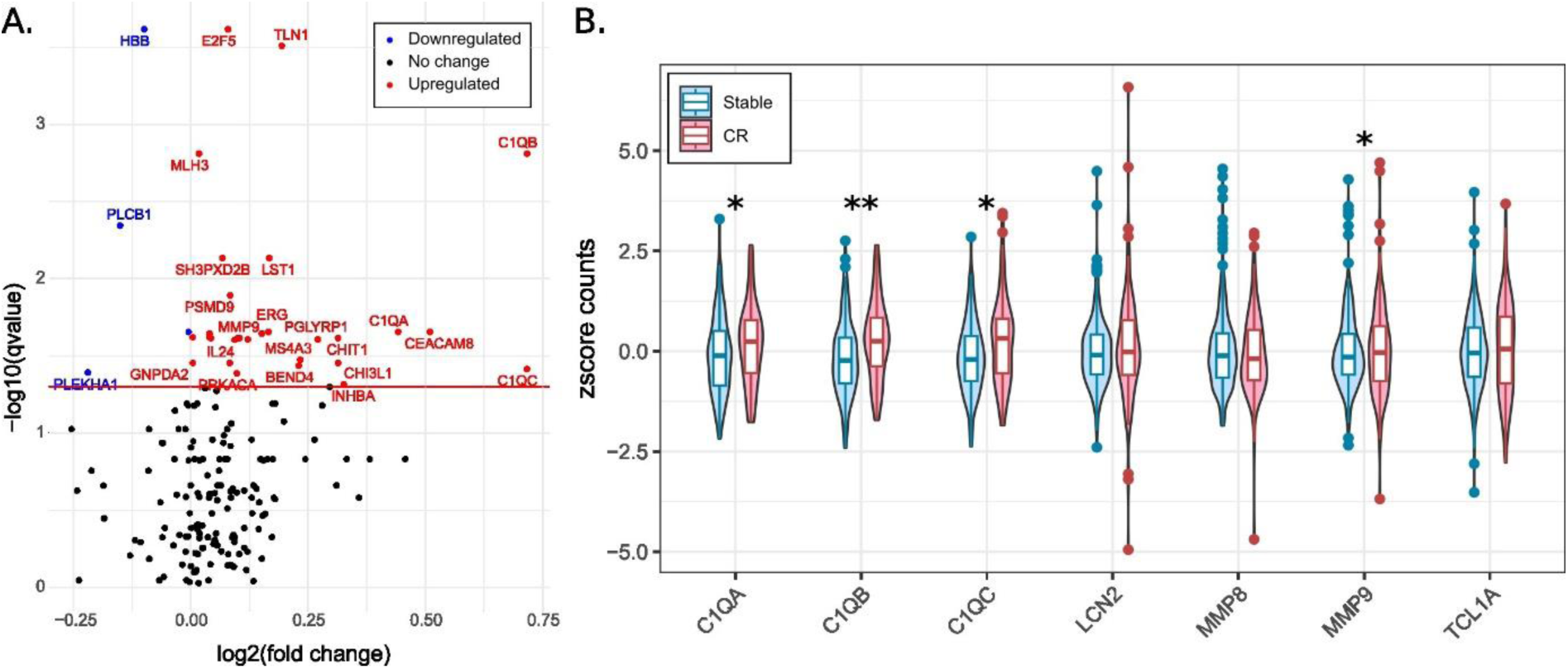
Meta-analysis of chronic rejection in six independent transcriptomic datasets. **A.** Volcano plot of the meta-analysis run in six independent cohorts for the 188 DEG identified in the discovery cohort. Genes colored in blue (4) and red (29) were considered significantly downregulated and upregulated respectively (qvalue ≤ 0.05). **B.** Violin plots representing global z-score expression of *C1QA*, *C1QB*, *C1QC*, *LCN2*, *MMP8*, *MMP9* and *TCL1A* in the six independent cohorts used in the meta-analysis. Expression values were normalized between cohorts in z-score in order to represent them globally. *qvalue ≤ 0.05, **qvalue ≤ 0.01 and ***qvalue≤0.001. *CR, chronic rejection*.

### MMP9 expression at 1-year post-transplant is downregulated in samples prior CAMR

To pinpoint early changes in PBMC gene expression associated with late subsequent CAMR, we collected 62 stable 1-year Reference patients without on-going or any previously reported rejection episode, including 11 who later experienced a CAMR event within 12.5 years. No significant difference was observed between the two subgroups for the demographics or clinical characteristics of the recipients, donors and graft follow-up during the first year, despite a trend for more *de novo* DSA reported in the post-1 year CAMR group (p=0.059, Table 3). The demographics and characteristics at the time of transplantation were also very comparable to the late CAMR/Stable initial groups (mostly male patients transplanted for the first time, with an average of 3 out of 6 *HLA* ABDR donor-recipient mismatches).

**Table 3:**
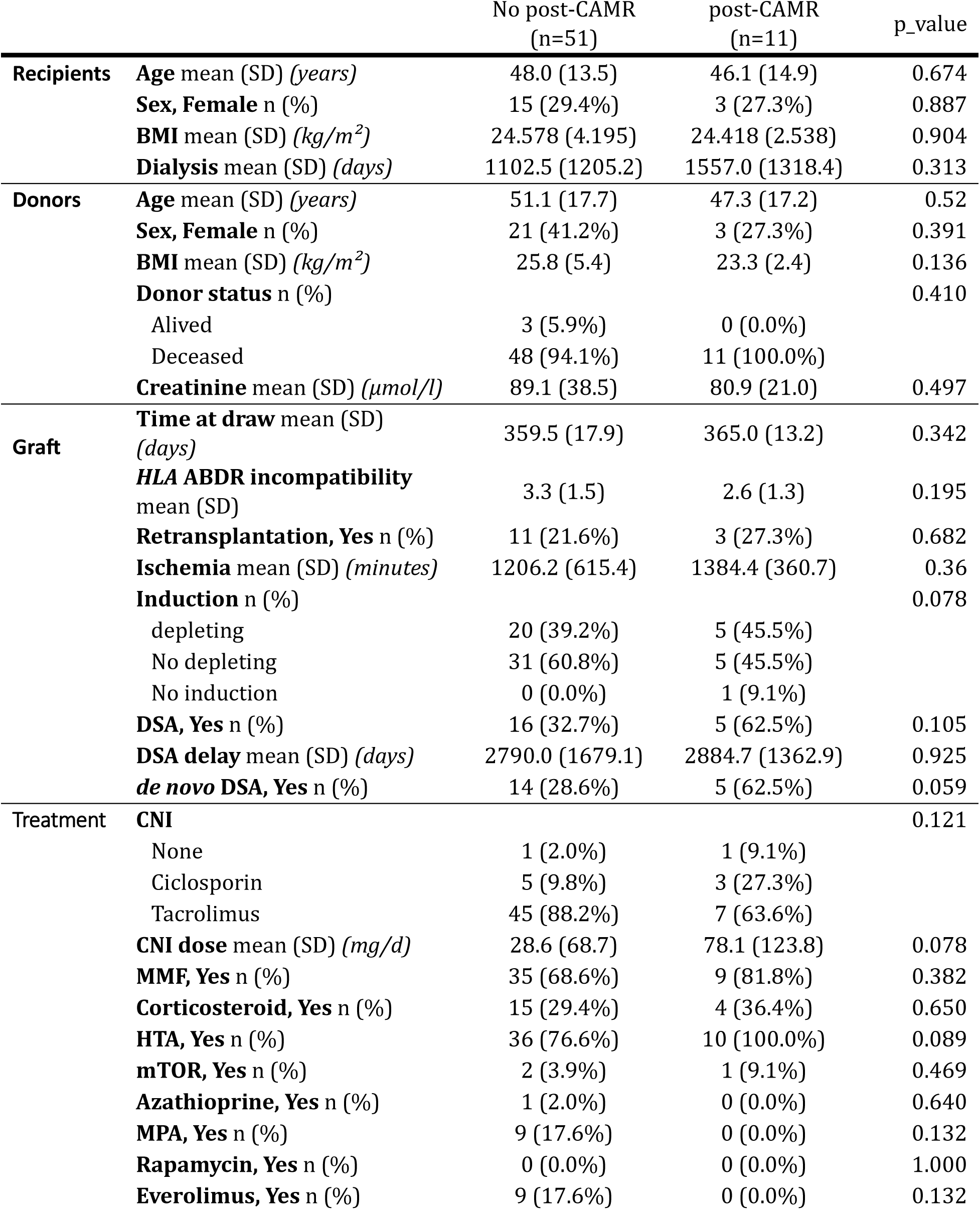
Characteristics of Stable 1-year Reference patients. Stable 1-year Reference individuals without any rejection event reported during the first-year post-transplantation were divided between patients who remained stable over subsequent follow-up (n=51) and those who later experienced CAMR event (within >1year and up to 12.5 years after transplantation, n=11). Categorical data (*e.g.* sex) are presented in absolute values and percentages, and were compared between both groups of interest with a chi-squared test. Continuous data (*e.g.* age) are presented as mean with standard deviation (SD), and were tested between both groups using an ANOVA t-test with equal variances.

The differential expression analysis between 1-year Reference patients who remained stable and those who later experienced a CAMR showed the significant upregulation of 3 genes and downregulation of 43 genes, including *MMP9* (log_2_(FC)=-3.38, q-value=1.27×10^-3^) (Figures 6A and 6B, Suppl. Table 6). The degranulation pathway was strongly enriched within the significantly downregulated genes at 1-year within PBMCs of kidney transplanted patients who will experience a CAMR (p=2.0×10^-11^), as well as for several immune regulation pathways (Suppl. Figure 5). Interestingly, 8 significant genes at 1-year were previously identified in our initial CAMR diagnosis signature, but exhibited opposite effect directions - being downregulated at 1-year in patients who later experienced CAMR, while being upregulated at CAMR diagnosis. Beyond *MMP9*, *CXCL1* (log_2_(FC)=-3.66, q-value=1.27×10^-3^), *MGAM2* (log_2_(FC)=-3.42, q-value=6.24×10^-3^), *BMX* (log_2_(FC)=-3.49, q-value=6.24×10^-3^), *MGAM* (log_2_(FC)=-2.45, q-value=9.39×10^-3^), *CYP4F3* (log_2_(FC)=-2.49, q-value=9.39×10^-3^), *KCNJ15* (log_2_(FC)=-2.35, q-value=1.66×10^-2^), and *PGLYRP1* (log_2_(FC)=-2.42, q-value=2.48×10^-2^) were downregulated at 1-year in patients who later presented with CAMR (Figure 5A). These 8 genes are all actors of the immune response regulation, including humoral responses, chemokine signaling and degranulation. These findings shed light on potential early biomarkers of CAMR by identifying gene expression alterations even before the onset of clinical manifestations.

**Figure 6:**
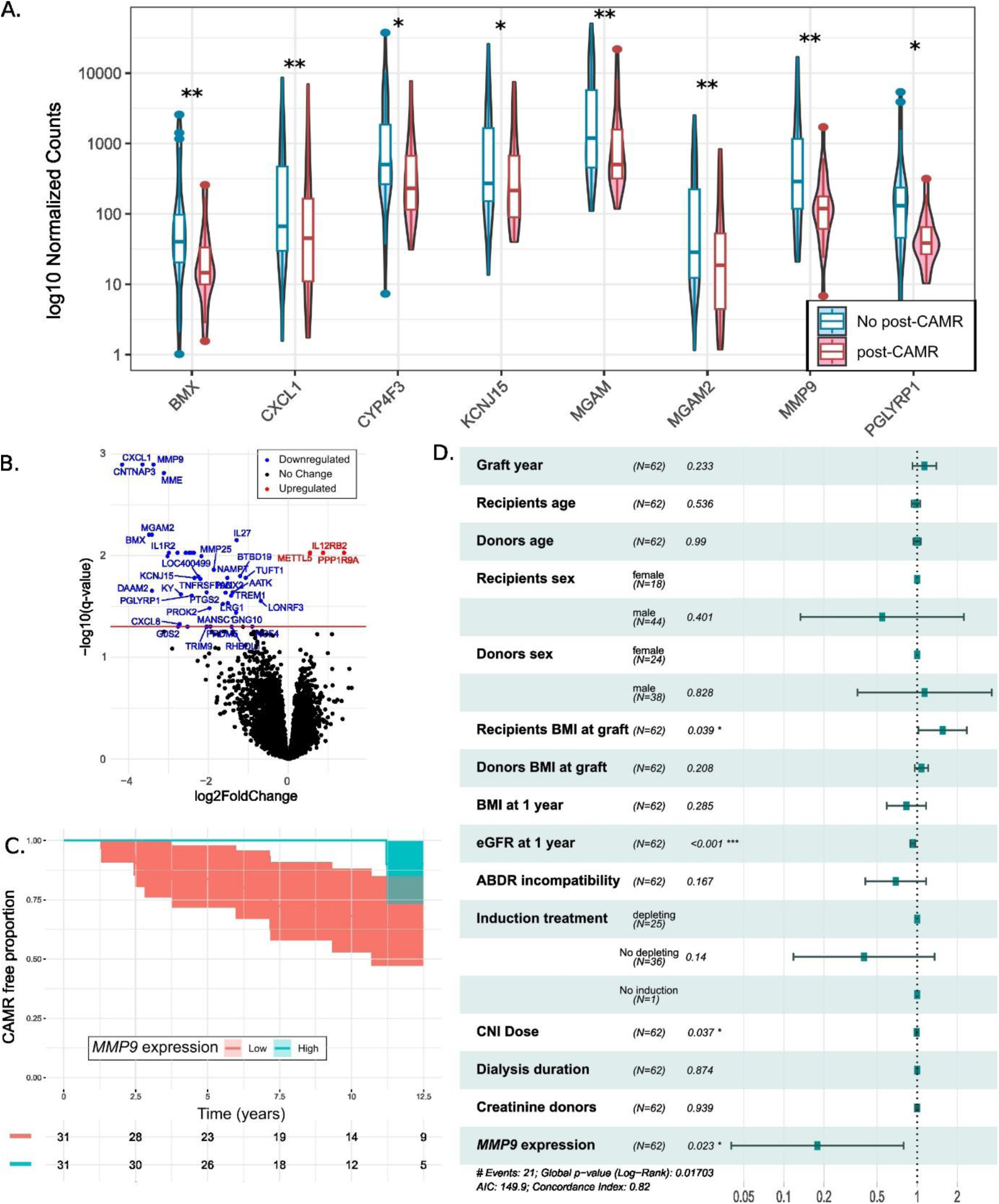
Impact of 1-year MMP9 expression on later CAMR occurrence. **A.** Violin plots showing PBMC gene expression for 1-year stable reference patients who remained stable (blue) and those who later experienced a post 1-year CAMR event (red). *BMX, CXCL1, CYP4F3, KCNJ15, MGAM, MGAM2, MMP9* and *PGLYRP1* are among the most significant genes and were associated with our CAMR diagnosis signature. **B.** Volcano plot showing the summary statistics of the differential expression analysis between 1-year stable reference patients who remained stable and those who later experienced a post 1-year CAMR event. Downregulated (43) and upregulated (3) genes are colored in blue and red respectively (qvalue ≤ 0.05). **C.** Kaplan-Meier plot depicting the CAMR-free proportion at 12.5 years based on high (blue) or low (red) *MMP9* expression (according to *MMP9* median expression) at 1-year post-transplantation. **D.** Forest plot showing the Cox model results for different variables collected at the time of graft and at 1-year. eGFR at 1-year, CNI dose at 1-year and *MMP9* are significant to predict a CAMR event before 12.5 year post-transplantation. *p≤0.05, **p≤0.01, and ***p≤0.001.

Finally, by stratifying patients into two groups based on their *MMP9* expression levels at 1-year post-transplantation, we observed that low *MMP9* 1-year expression was associated with an increased risk for developing CAMR over time (Figure 6C). We further implemented a multivariable Cox model to estimate the influence of several graft features in addition from *MMP9* 1-year expression on time-to-CAMR outcome (Figure 6D). Remarkably, higher eGFR (HR=0.93, p<0.001), high *MMP9* expression (HR=0.18, p=0.023), and higher CNI dose (HR=0.99, p=0.037) at 1 year were significant protective predictors of >1-year CAMR (Figure 6D). On the opposite, a higher recipient’s BMI at the time of graft was a risk factor for late CAMR (HR=1.55, p=0.039).

## Discussion

In the past decades, the number of patients on waiting lists for a kidney transplant has increased faster than the number of available grafts. CAMR is the primary cause of late kidney graft failure, and improving its diagnosis or prediction is therefore central to tackle allograft loss and limit retransplantations. In this study, we investigated PBMC gene expression patterns associated with CAMR. At the time of CAMR diagnosis, we found an upregulation of genes linked to the degranulation/ extracellular matrix pathway (including *MMP9*, *MMP8*, *LCN2*, and *CAMP*) and to the C1q complement complex (*C1QA*, *C1QB* and *C1QC*), while negative regulators of immune responses (*e.g. TCL1A*, *CXCR5*, *IL24* and *TRAF5*) were downregulated.

Importantly, we validated the *C1QA*, *C1QB*, *C1QC* and *MMP9* upregulation during chronic rejection in a meta-analysis of 6 independent transcriptomic datasets, including the only previous CAMR study^4^, underlying the robustness of our study design. The highlighted genes represent priority candidates for further disentangling the CAMR pathophysiology with functional assays and could be investigated as potential diagnosis biomarkers or drug targets. The meta-analysis did not confirm the *TCL1A* decreased peripheral expression during CAMR, which might be due to study design differences between our cohort and the replication datasets. However, the relevance of *TCL1A* in kidney transplantation has previously repeatedly been emphasized. Indeed, *TCL1A* blood expression was negatively associated with subclinical rejection at one-year post-transplantation and was validated as a biomarker^6^. Additionally, *TCL1A* blood expression has been shown to be negatively associated with acute rejection and DSA production^16,17^, and positively associated with operational tolerance in kidney transplantation^18,19^. *TCL1A* is predominantly expressed in naive B cells, as observed in our scRNA-seq analysis, highlighting their central role in graft outcomes such as rejection or operational tolerance^10,20^. Our findings hence confirmed the importance of *TCL1A* expression over the course of kidney transplantation, with a gradient of expression ranging from highest expression levels in operational tolerant patients, to stable patients, subclinical rejection, and finally to low expression in CAMR patients.

Our analysis focused on CAMR gene expression profiles and we assessed associations with biopsy lesions in order to gain insight on the rejection subtype specificity of our top candidates. As expected, the degranulation, complement, and *TCL1A* genes were strongly associated with C4d deposits and cg lesions, which are both hallmarks of CAMR. Interestingly, *C1QB* and *TCL1A* were also associated with t and i lesions, which are TCMR markers. This finding underlines the fact that some of the identified genes would be specific of the AMR/CAMR rejection subtypes, while others could play a broader role during immune responses associated with rejection. Investigating the expression profiles of these genes in additional samples exhibiting different rejection phenotypes will be necessary to further define their action mechanisms.

We have observed and validated an upregulation of C1q complement complex genes during CAMR. The C1q complex plays a crucial role in immune response initiation against the graft by binding to antibodies (including DSA) and activating the classical complement cascade^21^. Van Loon *et al.* reported an overexpression of *C1QA* both in biopsies and blood samples during rejection events and of *C1QB* specifically during CAMR^2^. This previous report strengthens our results and underlines the relevance of the peripheral compartment for investigating rejection events. It also emphasizes the complexity of discriminating molecular mechanisms specific to each rejection subtype, since *C1QA* was reported across rejection phenotypes in their study but was not associated with t or i lesions in our own study, and *C1QB* was reported as specific of CAMR in their study but also showed associations with t and i lesions in our study. Conflicting results regarding the importance of C1q binding to antibodies in graft survival exist. Some studies suggest that, in the presence of DSA, C1q is associated with a worse graft survival^22,23^, while other studies found a neutral effect^24,25^. Finally, scRNA-seq data from kidney-transplanted patients revealed the expression of C1q complex genes in CD16^+^ non-classical monocytes. Allograft infiltration of monocytes was already pointed out as a potential marker of transplant rejection^26^, and single cell transcriptomics highlighted the association between inflammation severity and monocytes infiltration in the allograft^27^, confirming their key role for tissue injury.

Finally, we ran a preliminary analysis to describe possible early predictive biomarkers of late CAMR. As for the signature at CAMR diagnosis, the degranulation and additional immune regulation pathways were also enriched at 1-year in the periphery of patients who subsequently experienced a CAMR event. Interestingly, 8 genes, including *MMP9*, were previously identified in our initial CAMR diagnosis analysis, but with opposite effect directions. *MMP9* was downregulated at 1-year in patients who later experienced CAMR, while it was upregulated at the time of CAMR diagnosis, suggesting a major and dynamic role for *MMP9* over the kidney transplantation course. The C1q complement genes and *TCL1A* expression did not significantly differ in our 1-year stable Reference samples. *MMP9* is a soluble metalloproteinase known to be involved in developmental processes, tissue remodelling, inflammatory responses and proliferative signaling pathways. *MMP9* plays a role in kidney development^28^ and kidney diseases, more specifically in acute kidney injury (AKI) and kidney fibrosis. Interestingly, *MMP9* involvement in kidney diseases appears to be ambiguous^29^. Indeed, it seems protective against AKI by its action on fibrin degradation, as evidenced in *mmp9* knockout mice models^30–32^. However, *MMP9* seems to have a pathological role in the AKI to CKD transition and increases macrophage and neutrophil infiltration in AKI mice models^30,33–35^. MMP9 major partner, NGAL (encoded by the *LCN2* gene), is increased 300-fold in blood and 1000-fold in urine during AKI, and is suggested as a potential biomarker of AKI^36,37^. These reports are consistent with the dual *MMP9* roles highlighted in the present CAMR study, with an early suggested protective effect for high *MMP9* levels that later turn into a marker of rejection. It is worth noting that we also observed an increase of *LCN2* expression at the time of CAMR. In lung transplantation, *MMP9* is known to be associated with bronchiolitis obliterans syndrome and chronic lung allograft dysfunction and was suggested as a potential biomarker^38–40^. Plasmatic levels of active MMP9 are increased at 7 days and 14 days post-lung transplantation before decreasing at 1 month without reaching the pre-transplantation level (n=46)^41^. MMP9-associated genes such as *MMP8* or *BPI* were shown to be upregulated at respectively 1 week and 3 months in kidney-transplanted patients’ blood, suggesting an early involvement of this pathway in kidney transplantation^42^. Together, these data suggest that *MMP9* kinetics may play a crucial role over the course of kidney transplantation and in CAMR development. *MMP9* peripheral expression therefore appears as a promising biomarker candidate for kidney transplantation follow-up, and looking at early (*e.g.* 1 month) and late (*e.g.* > 2 years) timepoints to refine the dynamic description of *MMP9* expression may be important to decipher MMP9 effects in kidney-transplanted patients. Additionally, functional analyses will be necessary to understand the pathophysiological molecular mechanisms of MMP9 during kidney transplantation. MMP9 role on the extracellular matrix remodelling may be critical in the first year of transplantation to clear early excess of fibrin and repair ischemia reperfusion injuries in the graft^43,44^. On the other hand, if persistent over time, MMP9 could later promote increased microvascular leakage and favor infiltration of immune cells, hence fostering chronic inflammation and renal graft injury^45,46^. In this line, we observed that *MMP9* expression is increased during severe chronic glomerulitis lesions (cg3, Figure 3B) -and not during lower grade cg lesions, reinforcing a detrimental role at later stages, when monocytes and neutrophils are recruited in the graft.

Despite our efforts, our report presents some limitations. First, even if we included one of the largest published monocentric cohorts with transcriptomic data on CAMR, the sample size might limit our statistical power for discovery and we could have missed CAMR genes with lower effect sizes. In particular for the predictive exploration within 1-year stable Reference samples, only 11 patients out of 62 later experienced CAMR. In addition, we did not have access to a replication cohort for our 1-year predictive signature, and these preliminary findings therefore require further validation. Second, we investigated the PBMC compartment, which is composed of a mixture of different cell populations. We addressed this limitation by evaluating single-cell data and performing a cell-type specific deconvolution within cell superpopulations. Our results could be refined by investigating cell subtypes and different cell activation and differentiation status.

In conclusion, our study has shed light on several crucial components of late CAMR. In particular, we underlined the paradoxical associations of high *MMP9* expression over time and context of tissue injury, with early protective association and later deleterious association at the time of diagnosis.Overall, the targets identified here represent priority candidates for further digging the CAMR pathophysiology mechanisms and investigating their potential as biomarkers or drug targets.

## Data Availability

All data produced in the present study are available upon reasonable request to the corresponding author.

## Data Sharing Statement

Our analytic pipeline is available at the following repository: https://gitlab.univ-nantes.fr/morin-m-2/imokit.

Raw data are available on the European Bioinformatics Institute (EBI) Annotare server at the accession numbers: E-MTAB-16340 (CAMR and Stable samples), E-MTAB-16370 (1-year samples).

## Acknowledgments

We thank everyone who helped in the design, collection, experiments, data cleaning and analyses. We are especially indebted to all recipients who participated in this study, to the clinical staff and physicians who care and manage the patients on a daily basis and helped recruiting the patients, and to research associates who participated in the data collection. The authors would like to thank the members of the DIVAT consortium for their involvement in the study, the physicians who helped recruit patients, and all patients who participated in this study. We also thank the clinical research associates who participated in the data collection. The analysis and interpretation of these data are the responsibility of the authors.

DIVAT (Données Informatisées et VAlidées en Transplantation) Consortium:

Nantes : Gilles Blancho, Julien Branchereau, Diego Cantarovich, Agnès Chapelet, Jacques Dantal, Clément Deltombe, Lucile Figueres, Claire Garandeau, Magali Giral, Caroline Gourraud-Vercel, Eloise Grenon, Clarisse Kerleau, Thibault Letellier, Christophe Masset, Aurélie Meurette, Simon Ville, Christine Kandell, Karine Renaudin, Florent Delbos, Alexandre Walencik, Anne Devis.

We thank the biological resource centre for biobanking (CHU Nantes, Nantes Université, Centre de ressources biologiques (BB-0033-00040), F-44093 Nantes, France). We are also grateful to the Bioinformatics Core Facility BiRD, member of Biogenouest and Institut Français de Bioinformatique (IFB) (ANR-11-INBS-0013) for the use of their resources and their technical support. We also thank Marwan Touati, Dr Valentin Quiniou and Parean Biotechnologies for their assistance with RNA-seq library preparations and sequencing as well as for their technical assistance in the analytical processes.

## Author Contributions

MM and LS conceived and designed the study. MM, KC, RK, GM and LS performed sample selection. GM, MC, VS, BG and GM monitored the patients in the clinics and shared their clinical expertise for study design and data interpretation. MM and DA performed the RNA sample extraction. MM ran all QC, bioinformatic and statistical analyses. BA, DR, BS, DA, VN shared their methodological expertise in bulk and/or single-cell transcriptomics, data analysis and integration. MM, MV and RO performed the Cox regression models and time-dependent analyses. LS supervised the project and provided critical revisions. All authors have read and approved the final version of the manuscript.

