## Supplementary material for "Peripheral transcriptomic signature of chronic antibody-mediated rejection in kidney transplantation: a dual effect for *MMP9* over time": Supllemental Material

Corresponding author information

Pr Sophie Limou, PhD

CHU Nantes Hôtel Dieu - ITUN - CR2TI

30 bd Jean Monnet

F-44093 Nantes cedex 1 ; France

#### Supplementary methods

##### *Sample selection and RNA extraction*

For the CAMR group, inclusion criteria were patients with biopsy-proven CAMR, with more than one year follow-up at the time at diagnosis and without any preceding recorded rejection (Figure 1). Biopsy-proven rejection was defined according to the Banff 2022 classification with the three following criteria: evidence of chronic tissue injury (indicated by cg>0 and/or severe ptcml), evidence of current/recent antibody interaction with the endothelium (indicated by C4d staining and/or moderate microvascular inflammation [(g+ptc)≥2]), and serologic evidence of circulating DSA (or C4d staining). The Stable group included patients with a stable graft function under immunosuppressive treatment, with more than one year follow-up at the time of biopsy evaluation, and with no current or preceding recorded rejection event (Figure 1). Our first transcriptomic analysis compared these CAMR vs. Stable groups to define the peripheral signature at the time of CAMR diagnosis.

The Reference group gathered patients with a stable graft function at 1-year post-transplantation and with no biopsy-proven rejection during the first-year post-transplantation (Figure 1). Within this group, 11 patients subsequently experienced a CAMR event (between >1-year and 12.5 years post-transplantation), and 51 patients had no recorded CAMR event during the same laps of follow-up time. Our second transcriptomic analysis compared 1-year stable patients who later experienced a CAMR vs. those who did not, in order to define an early subclinical CAMR signature and identify potential predictive biomarkers of late CAMR.

For all samples, PBMC samples were stored à -80°C in Trizol reagent (Thermo Fisher Scientific, Waltham USA), and we isolated RNA samples using the Direct-zol DNA/RNA miniprep kit (Zymo research, Seattle USA) according to the manufacturer instructions. RNA concentration and quality were assessed using the Nanodrop Spectrophotometer and Caliper electrophoresis (Perkin Elmer, Wellesley USA). All samples met the quality criteria of RNA integrity number above 8.0 and concentration above 30 ng/µL.

##### *Total RNA-seq library preparation*

Library preparations were outsourced and performed by Parean biotechnologies (Saint-Malo, France). Libraries were prepared using SMARTer Stranded Total RNA Sample Prep (Takara Bio, Mountain View USA) in accordance with the manufacturer guidelines, employing an ACSIA automate from Primadiag. The quality and validation of these libraries were assessed using the Tapestation 4200 system from HS D1000 analysis. The sequencing process entailed a minimum of 50 million reads per sample, employing paired-ended sequencing (2*100pb) on a Novaseq 6000 instrument (Illumina, San Diego CA, USA). Throughout this process, four sequencing lines were used to ensure optimal throughput and efficiency.

##### *Total RNA-seq differential expression analysis*

The sequencing fastq files underwent quality assessment with the fastqc tool (version 0.11.9) and were trimmed with trimmomatic (version 0.39) using the following criteria: leading and trailing base qualities >28, first 4 5' bases with a mean quality of 15, and reads >30 bp. Qualities were coded in phred33 scores. Alignment was performed using STAR (version 2.7.10a), followed by quality analysis of aligned reads using samtools (version 1.13) and deeptools (version 3.5.1). Gene expression quantification was conducted via featurecount (version 2.0.1) utilizing the refseq database (version 2021-02-10) sourced from NCBI annotations[1]. Normalization and differential expression analysis were carried out using RUVseq[2] (version 1.32.0) and DESeq2 R packages[3] (version 1.38.0), respectively.

All transcriptomic analyses were performed using generalized mixed models implemented in DESeq2 and corrected for patients' sex, age and BMI at sampling, post-transplantation time at sampling, sequencing line, and graft year. Differentially expressed genes were defined by a q-value (false discovery rate, FDR) below 0.05. Two missing BMI values were imputed using the mice R package (version 3.16.0). Pathway enrichment analyses were performed using metascape[4], which queries a large variety of databases (including gene ontology).

*Single-cell RNA-seq analysis*

The Seurat tool[5] (version 4.1.1) was used for the analysis of the GSE230651 scRNA-seq dataset. Classical quality control steps were implemented, including filtering cells with RNA counts ranging from 250 to 3000, and mitochondrial RNA below 10%. Clustering, Uniform Manifold Approximation and Projection (UMAP), and cell annotation were performed using azimuth (version 0.4.6) and the human PBMC reference panel from the HuBMAP consortium[6]. Cells with an azimuth prediction score below 0.7 or categorized as "other" were excluded from subsequent analyses.

*Deconvolution and synthetic scRNA-seq Analysis*

The signature matrix was built from the annotated GSE230651 scRNA-seq data using the recommended CIBERSORTx parameters, including quantile normalization and no filtration of non-hematopoietic genes. The cell fraction imputation was performed by enabling the following parameters: S-mode batch correction, quantile normalization, and relative mode, and employing 500 permutations. Subsequently, the cell expression imputation was performed by enabling the S-mode batch correction and quantile normalization parameters, and with a window size for deconvolution set to 28. Due to computation limitations, the cell-specific expression was imputed only for the top 1000 differentially expressed genes from the CAMR vs. Stable analysis.

*External validation of the CAMR diagnosis signature*

For each of the 6 transcriptomic datasets (see Table 1), we only extracted chronic rejection (CR, n=131) and stable (n=360) samples. CR samples from the GSE14655 dataset were biopsy-proven chronic rejection (n=35), while CR samples from the 5 remaining datasets were biopsy-proven CAMR (n=96). Using these independent datasets, we performed a meta-analysis on the 188 genes found differentially expressed during CAMR in the discovery cohort using the DExMA R package (1.14.0)[7] while accounting for the dataset and the technology (RNA-seq or microarray). As no individual clinical data were available for most studies, we could not correct the meta-analysis for additional covariates. We only retained genes with data available in ≥3 studies and patients with <33% of missing data, leaving 176/188 (94%)

genes and all patients. The Stouffer method with an FDR threshold of 0.05 was used to identify differentially expressed genes in this meta-analysis.

### Supplementary Figures

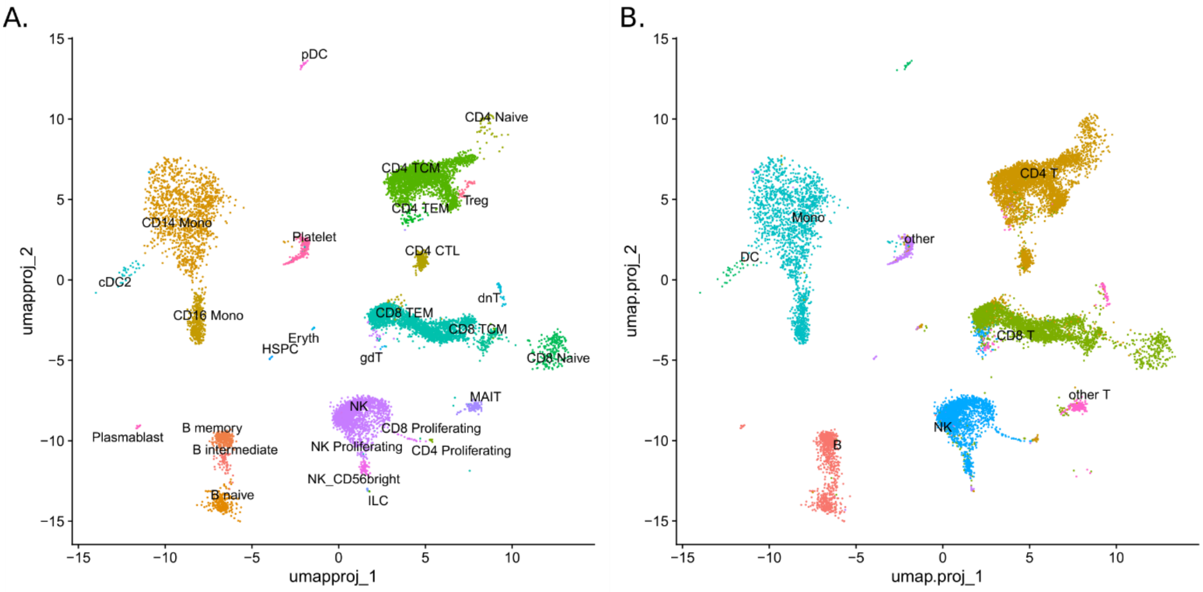


*Suppl. Figure 1: Cell clusters annotation from scRNA-seq data of kidney transplanted patients*

**A.** UMAP of kidney recipients' scRNAseq data annotated with the Azimuth PBMC reference dataset. This data is used in Figures 2A and 2B to explore the cell-specific expression of *TCL1A*, *C1QA* and *MMP9*. **B.** UMAP showing the same data annotated at a lower resolution to facilitate cell deconvolution and synthetic scRNA-seq of our study's bulk RNA-seq data.

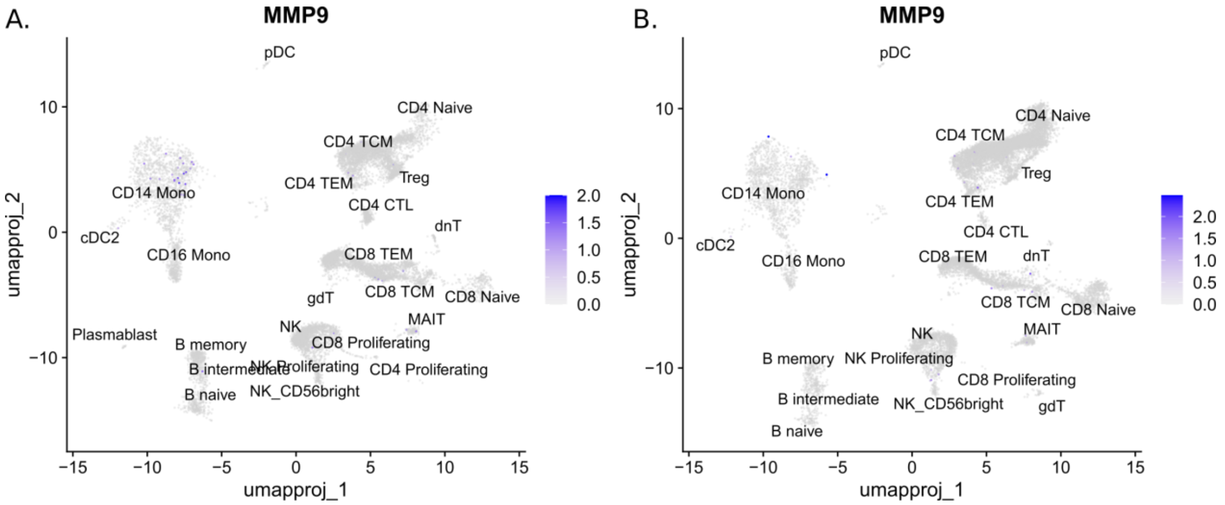


*Suppl. Figure 2: MMP9 is mainly expressed in CD14+ monocytes of kidney recipients*

**A.** UMAP of kidney recipients' scRNAseq data annotated with the Azimuth PBMC reference dataset. Cells expressing *MMP9* are colored in blue. **B.** UMAP of healthy individuals' scRNAseq data (GSE230651), showing that nearly no cells expressed *MMP9* in healthy individuals.

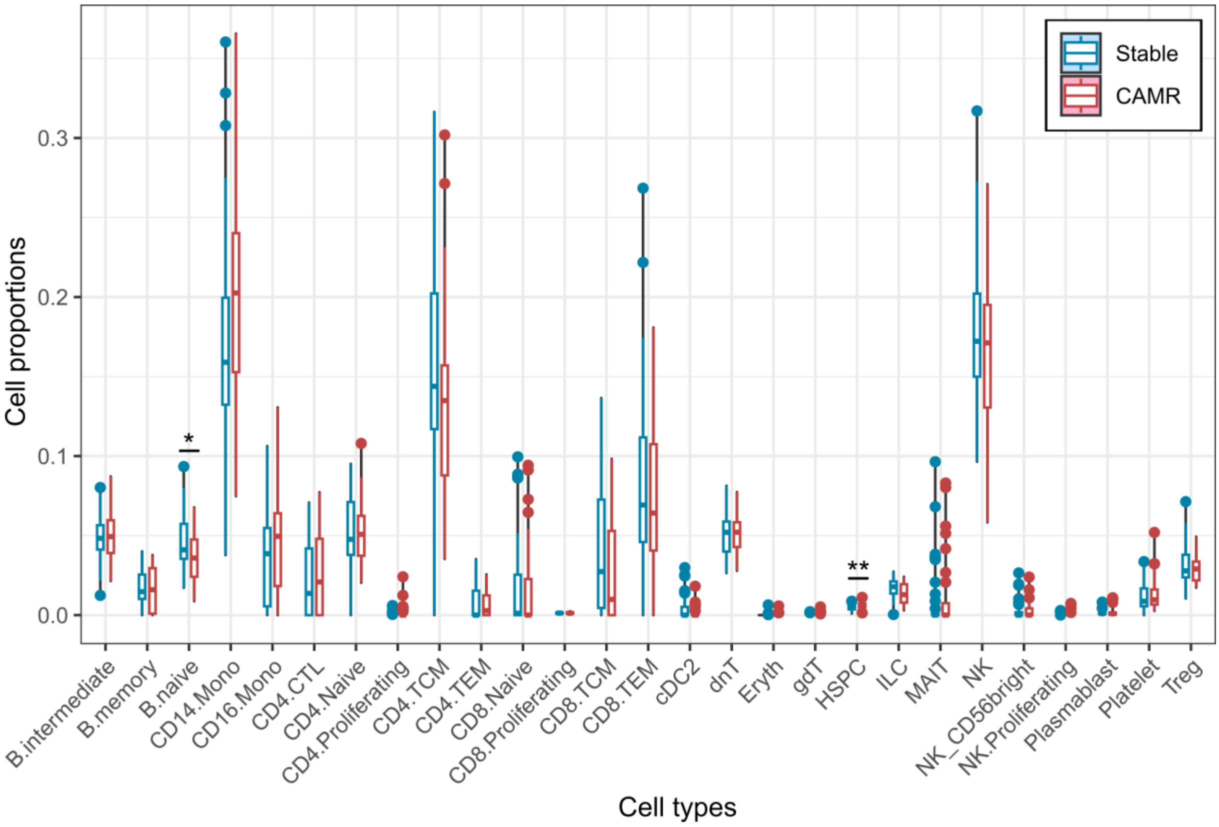


*Suppl. Figure 3: Imputed PBMC cell type proportions in our late Stable and CAMR patients*

Box plots presenting the imputed cell proportions in our Stable (blue) and CAMR (red) samples. No cell type was significantly different between both groups when accounting for multiple testing (Bonferroni threshold $p=1.9x10^{-3}$). Naive B cell and HSPC proportions were respectively nominally decreased and increased in CAMR samples. *p value≤0.05, and **p value≤0.01.

*Mono, monocyte; MAIT, mucosal associated invariant T cell; ILC, Innate lymphoid cell; Eryth, erythrocyte; TEM, T effector memory cell; cDC2, classical dendritic cell 2; CTL, cytotoxic T lymphocyte; HSPC, hematopoietic stem and progenitor cell; gdT, gamma-delta T cell; dnT, double negative T cell.*

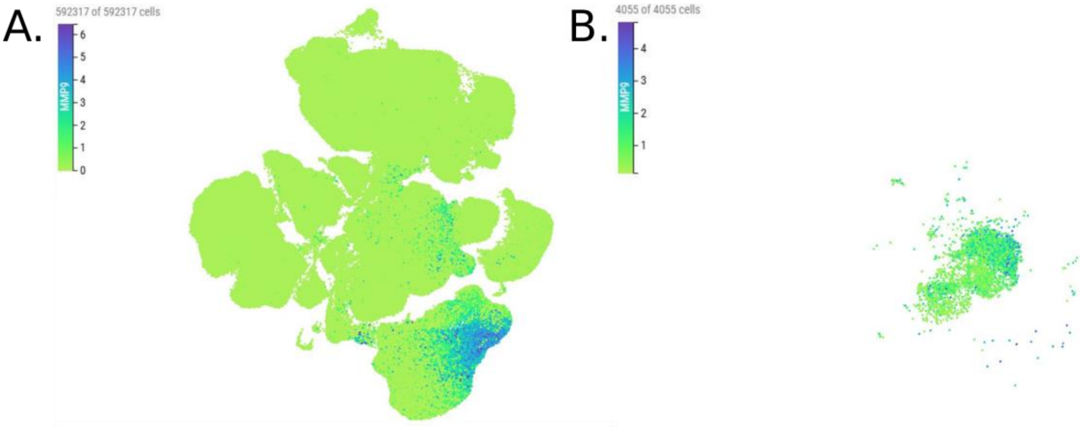


*Suppl. Figure 4: MMP9 expression in immune cells and monocytes*

A. *MMP9* expression is represented in all immune cells from the tabula sapiens consortium. 592,317 cells are represented in this figure representing immune cells from all body tissues analysed by the tabula sapiens consortium. Cells were colored with a gradient from green corresponding to low level of *MMP9* expression to blue corresponding to high level of *MMP9* expression.  B. Same dataset as A with filters for monocytes, classical monocytes and non-classical monocytes expressing *MMP9*. A total of 4,055 monocytes express *MMP9* in these data (out of 45,237 monocytes in total, *i.e.* 9%).

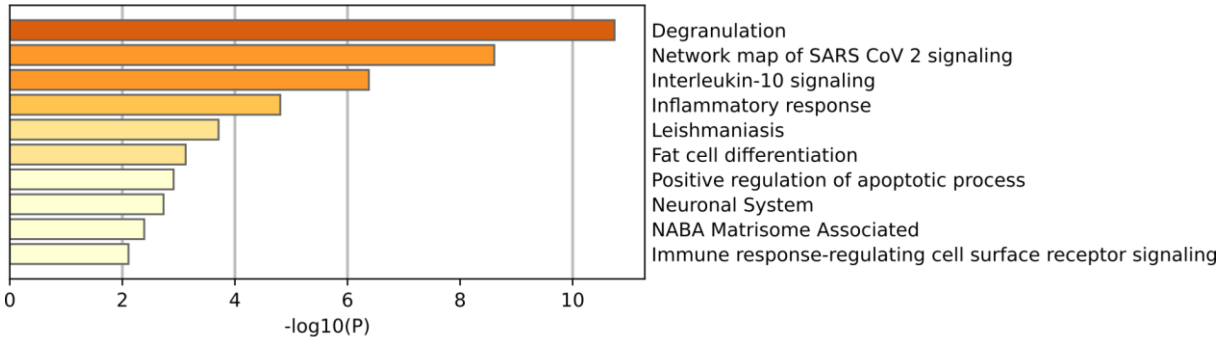


*Suppl. Figure 5: Pathway enrichment analysis of the significantly downregulated genes in 1-year stable Reference patients who later experienced a CAMR event*

Dark orange, orange and light orange bars reflect -$\log_{10}$(p-value) ≥10, ≥ 6, and ≥ 3, respectively.

| Gene | baseMean | log2FoldChange | lfcSE | stat | pvalue | qvalue |
|---|---|---|---|---|---|---|
| **MMP9** | 710.86 | 2.55 | 0.42 | 6.10 | 1.08E-09 | 1.36E-05 |
| **ANXA3** | 580.31 | 1.82 | 0.36 | 5.02 | 5.21E-07 | 0.00 |
| **BMX** | 90.40 | 2.65 | 0.54 | 4.90 | 9.80E-07 | 0.00 |
| **MMP8** | 1594.04 | 2.06 | 0.43 | 4.81 | 1.52E-06 | 0.00 |
| **LTF** | 4100.59 | 1.83 | 0.39 | 4.74 | 2.18E-06 | 0.00 |
| **HBB** | 1285.90 | 2.45 | 0.52 | 4.72 | 2.38E-06 | 0.00 |
| **CHIT1** | 42.29 | 2.09 | 0.44 | 4.71 | 2.52E-06 | 0.00 |
| **PGLYRP1** | 199.88 | 1.79 | 0.38 | 4.68 | 2.81E-06 | 0.00 |
| **CXCR5** | 325.86 | -0.93 | 0.20 | -4.65 | 3.32E-06 | 0.00 |
| **ZNF154** | 412.79 | -0.65 | 0.14 | -4.61 | 3.93E-06 | 0.00 |
| **CSNK1A1L** | 23.20 | 2.27 | 0.49 | 4.61 | 4.01E-06 | NA |
| **ZNF814** | 486.25 | -0.44 | 0.10 | -4.60 | 4.32E-06 | 0.00 |
| **CEACAM6** | 422.93 | 1.92 | 0.43 | 4.46 | 8.31E-06 | 0.01 |
| **ABCA13** | 984.19 | 1.69 | 0.38 | 4.44 | 9.12E-06 | 0.01 |
| **CEACAM8** | 540.38 | 1.76 | 0.40 | 4.37 | 1.26E-05 | 0.01 |
| **SH3YL1** | 318.15 | -0.54 | 0.13 | -4.27 | 1.92E-05 | 0.02 |
| **TCL1A** | 154.55 | -1.26 | 0.29 | -4.27 | 1.95E-05 | 0.02 |
| **TRAF5** | 1220.85 | -0.37 | 0.09 | -4.25 | 2.18E-05 | 0.02 |
| **MAML3** | 1512.99 | 0.48 | 0.11 | 4.24 | 2.19E-05 | 0.02 |
| **C1QB** | 63.33 | 1.37 | 0.33 | 4.20 | 2.72E-05 | 0.02 |
| **ARRB2** | 7400.39 | 0.31 | 0.07 | 4.18 | 2.88E-05 | 0.02 |
| **ZNF84** | 533.78 | -0.37 | 0.09 | -4.17 | 3.07E-05 | 0.02 |
| **RIN3** | 4263.57 | 0.24 | 0.06 | 4.16 | 3.12E-05 | 0.02 |
| **CAMP** | 360.92 | 1.55 | 0.38 | 4.11 | 4.04E-05 | 0.02 |
| **NUDT16** | 1847.22 | 0.51 | 0.13 | 4.08 | 4.59E-05 | 0.02 |
| **ERG** | 97.19 | 1.38 | 0.34 | 4.07 | 4.61E-05 | 0.02 |
| **YWHAH** | 1151.80 | 0.41 | 0.10 | 4.07 | 4.74E-05 | 0.02 |
| **FLT3** | 527.89 | 0.89 | 0.22 | 4.06 | 4.89E-05 | 0.02 |
| **LIPT1** | 441.14 | -0.36 | 0.09 | -4.06 | 4.99E-05 | 0.02 |
| **ARHGEF40** | 2666.23 | 0.55 | 0.14 | 4.05 | 5.15E-05 | 0.02 |
| **BEND7** | 20.21 | 1.58 | 0.40 | 4.00 | 6.28E-05 | NA |
| **SH3PXD2B** | 202.63 | 0.95 | 0.24 | 4.00 | 6.45E-05 | 0.03 |
| **PLPP5** | 391.49 | -0.43 | 0.11 | -3.99 | 6.50E-05 | 0.03 |
| **HELQ** | 1007.12 | -0.27 | 0.07 | -3.99 | 6.70E-05 | 0.03 |
| **ZNF248** | 408.63 | -0.32 | 0.08 | -3.97 | 7.14E-05 | 0.03 |
| **MACIR** | 90.40 | 0.77 | 0.20 | 3.96 | 7.38E-05 | 0.03 |
| **TBXAS1** | 2775.98 | 0.46 | 0.12 | 3.95 | 7.91E-05 | 0.03 |
| **SPTB** | 132.98 | 1.11 | 0.28 | 3.94 | 8.04E-05 | 0.03 |
| **ZBTB25** | 1638.62 | -0.35 | 0.09 | -3.94 | 8.17E-05 | 0.03 |
| **ZFP14** | 462.93 | -0.33 | 0.08 | -3.93 | 8.50E-05 | 0.03 |
| **GNPNAT1** | 523.88 | -0.28 | 0.07 | -3.92 | 8.97E-05 | 0.03 |
| **SEMA4G** | 51.57 | -0.61 | 0.16 | -3.91 | 9.36E-05 | 0.03 |
| **FIGNL1** | 406.12 | -0.41 | 0.10 | -3.90 | 9.74E-05 | 0.03 |
| **KREMEN1** | 292.99 | 1.20 | 0.31 | 3.88 | 0.0001 | 0.03 |
| **SAP30** | 737.21 | 0.90 | 0.23 | 3.87 | 0.0001 | 0.03 |

| | | | | | | |
|---|---|---|---|---|---|---|
| **LRPAP1** | 1289.27 | 0.25 | 0.06 | 3.87 | 0.0001 | 0.03 |
| **FBXO22** | 651.16 | -0.29 | 0.08 | -3.86 | 0.0001 | 0.03 |
| **IL24** | 382.35 | -0.76 | 0.20 | -3.85 | 0.0001 | 0.03 |
| **TLN1** | 26216.38 | 0.28 | 0.07 | 3.84 | 0.0001 | 0.03 |
| **C1QA** | 157.13 | 0.97 | 0.25 | 3.84 | 0.0001 | 0.03 |
| **PSMD9** | 246.64 | 0.34 | 0.09 | 3.84 | 0.0001 | 0.03 |
| **BPI** | 1086.94 | 1.33 | 0.35 | 3.83 | 0.0001 | 0.03 |
| **CRISP3** | 518.14 | 1.52 | 0.40 | 3.83 | 0.0001 | 0.03 |
| **STAP1** | 297.95 | -0.69 | 0.18 | -3.81 | 0.0001 | 0.03 |
| **PLCB1** | 1646.23 | 0.43 | 0.11 | 3.81 | 0.0001 | 0.03 |
| **ORM1** | 59.98 | 1.20 | 0.31 | 3.81 | 0.0001 | 0.03 |
| **PCID2** | 898.18 | -0.27 | 0.07 | -3.79 | 0.0001 | 0.03 |
| **WDR89** | 708.29 | -0.35 | 0.09 | -3.79 | 0.0002 | 0.03 |
| **TNRC18** | 6574.62 | 0.27 | 0.07 | 3.78 | 0.0002 | 0.03 |
| **LCN2** | 198.86 | 1.41 | 0.37 | 3.78 | 0.0002 | 0.03 |
| **KCNG1** | 23.78 | -1.46 | 0.39 | -3.78 | 0.0002 | NA |
| **CEACAM1** | 465.15 | 1.14 | 0.30 | 3.78 | 0.0002 | 0.03 |
| **ZCCHC4** | 406.07 | -0.28 | 0.07 | -3.78 | 0.0002 | 0.03 |
| **GNPDA2** | 327.97 | -0.37 | 0.10 | -3.78 | 0.0002 | 0.03 |
| **APOBR** | 1053.80 | 0.61 | 0.16 | 3.77 | 0.0002 | 0.03 |
| **ERGIC1** | 2397.19 | 0.31 | 0.08 | 3.77 | 0.0002 | 0.03 |
| **BTLA** | 582.05 | -0.54 | 0.14 | -3.77 | 0.0002 | 0.03 |
| **PNPLA7** | 145.38 | -0.59 | 0.16 | -3.77 | 0.0002 | 0.03 |
| **LPL** | 43.33 | 1.54 | 0.41 | 3.77 | 0.0002 | 0.03 |
| **ARHGAP29** | 113.02 | 0.97 | 0.26 | 3.76 | 0.0002 | 0.03 |
| **ZBTB7B** | 3700.90 | 0.31 | 0.08 | 3.75 | 0.0002 | 0.03 |
| **E2F5** | 108.25 | -0.70 | 0.19 | -3.75 | 0.0002 | 0.03 |
| **G6PD** | 1377.95 | 0.40 | 0.11 | 3.75 | 0.0002 | 0.03 |
| **RGS19** | 1224.76 | 0.38 | 0.10 | 3.75 | 0.0002 | 0.03 |
| **TXN** | 335.62 | 0.41 | 0.11 | 3.74 | 0.0002 | 0.03 |
| **CEBPE** | 63.06 | 1.13 | 0.30 | 3.74 | 0.0002 | 0.03 |
| **TCN1** | 302.56 | 1.40 | 0.37 | 3.74 | 0.0002 | 0.03 |
| **STXBP2** | 1839.87 | 0.29 | 0.08 | 3.73 | 0.0002 | 0.03 |
| **CXCL1** | 336.75 | 1.71 | 0.46 | 3.73 | 0.0002 | 0.03 |
| **CLIC1** | 4462.58 | 0.32 | 0.09 | 3.73 | 0.0002 | 0.03 |
| **HTRA3** | 16.72 | 1.80 | 0.48 | 3.73 | 0.0002 | NA |
| **FAM111B** | 166.23 | -0.88 | 0.24 | -3.72 | 0.0002 | 0.03 |
| **CD163L1** | 23.16 | 1.20 | 0.32 | 3.72 | 0.0002 | NA |
| **MLH3** | 1112.62 | -0.24 | 0.06 | -3.72 | 0.0002 | 0.03 |
| **IARS1** | 1830.54 | -0.31 | 0.08 | -3.72 | 0.0002 | 0.03 |
| **HK1** | 5700.18 | 0.26 | 0.07 | 3.72 | 0.0002 | 0.03 |
| **JDP2** | 921.56 | 0.63 | 0.17 | 3.71 | 0.0002 | 0.03 |
| **INHBA** | 155.20 | 1.47 | 0.40 | 3.70 | 0.0002 | 0.03 |
| **CFL1** | 6775.82 | 0.24 | 0.06 | 3.70 | 0.0002 | 0.03 |
| **THOC3** | 177.91 | -0.41 | 0.11 | -3.70 | 0.0002 | 0.03 |
| **PLEKHA1** | 1235.97 | -0.33 | 0.09 | -3.70 | 0.0002 | 0.03 |

| | | | | | | |
|---|---|---|---|---|---|---|
| **FICD** | 278.42 | -0.36 | 0.10 | -3.70 | 0.0002 | 0.03 |
| **ZNF527** | 139.13 | -0.40 | 0.11 | -3.69 | 0.0002 | 0.03 |
| **ALG9** | 559.29 | -0.29 | 0.08 | -3.68 | 0.0002 | 0.03 |
| **TAPT1** | 1480.86 | -0.32 | 0.09 | -3.68 | 0.0002 | 0.03 |
| **OSBP2** | 141.70 | 0.72 | 0.20 | 3.68 | 0.0002 | 0.03 |
| **CRISPLD2** | 1625.46 | 0.63 | 0.17 | 3.67 | 0.0002 | 0.03 |
| **PPP4C** | 1212.52 | 0.41 | 0.11 | 3.67 | 0.0002 | 0.03 |
| **TSPAN16** | 124.47 | 0.82 | 0.22 | 3.67 | 0.0002 | 0.03 |
| **PILRA** | 1513.60 | 0.44 | 0.12 | 3.66 | 0.0003 | 0.03 |

*Suppl. Table 1: Summary statistics of the top 100 DEG between Stable (n=43) vs. CAMR (n=35) patients*

*baseMean, base mean expression; log2FoldChange, log2 transformation of the fold change between CAMR and Stable patients; lfcSE, log2(Fold change) standard error;* stat, Wald statistic.

| GroupID | Category | Term | Description | LogP | Log(q-value) |
|---|---|---|---|---|---|
| 1_Summary | Reactome Gene Sets | R-HSA-6798695 | Neutrophil degranulation | -20.6 | -16.3 |
| 2_Summary | KEGG Pathway | hsa05168 | Herpes simplex virus 1 infection | -7.6 | -3.7 |
| 3_Summary | GO Biological Processes | GO:0002683 | negative regulation of immune system process | -7.5 | -3.7 |
| 4_Summary | GO Biological Processes | GO:0071396 | cellular response to lipid | -7.4 | -3.6 |
| 5_Summary | CORUM | CORUM:6418 | C1q complex | -6.6 | -3.0 |
| 6_Summary | GO Biological Processes | GO:1902105 | regulation of leukocyte differentiation | -6.1 | -2.5 |
| 7_Summary | GO Biological Processes | GO:0002697 | regulation of immune effector process | -6.0 | -2.5 |
| 8_Summary | GO Biological Processes | GO:0032680 | regulation of tumor necrosis factor production | -5.6 | -2.2 |
| 9_Summary | KEGG Pathway | hsa04062 | Chemokine signaling pathway | -5.5 | -2.2 |
| 10_Summary | Reactome Gene Sets | R-HSA-1566977 | Fibronectin matrix formation | -5.3 | -2.1 |
| 11_Summary | Canonical Pathways | M219 | PID HEDGEHOG GLI PATHWAY | -4.9 | -1.9 |
| 12_Summary | GO Biological Processes | GO:0001775 | cell activation | -4.9 | -1.9 |
| 13_Summary | GO Biological Processes | GO:0006959 | humoral immune response | -4.8 | -1.9 |
| 14_Summary | GO Biological Processes | GO:0071345 | cellular response to cytokine stimulus | -4.8 | -1.9 |
| 15_Summary | GO Biological Processes | GO:1901072 | glucosamine-containing compound catabolic process | -4.7 | -1.8 |
| 16_Summary | GO Biological Processes | GO:0001934 | positive regulation of protein phosphorylation | -4.5 | -1.7 |
| 17_Summary | Reactome Gene Sets | R-HSA-109582 | Hemostasis | -4.4 | -1.7 |
| 18_Summary | GO Biological Processes | GO:0010569 | regulation of double-strand break repair via homologous recombination | -4.0 | -1.4 |
| 19_Summary | Reactome Gene Sets | R-HSA-5674135 | MAP2K and MAPK activation | -4.0 | -1.4 |
| 20_Summary | GO Biological Processes | GO:0060627 | regulation of vesicle-mediated transport | -3.9 | -1.3 |

*Suppl. Table 2: Pathway enrichment analysis of the DEG between Stable and CAMR patients*

This table presents the summary statistics of the pathway enrichment analysis run from the DEG between Stable and CAMR patients using Metascape.

| | Stable (N=43) | CAMR (N=35) | p value |
|---|---|---|---|
| **CD4 T** | | | 0.275 |
| Mean (SD) | 0.358 (0.133) | 0.326 (0.118) | |
| Range | 0.094 - 0.643 | 0.116 - 0.623 | |
| **CD8 T** | | | 0.945 |
| Mean (SD) | 0.122 (0.098) | 0.121 (0.085) | |
| Range | 0.000 - 0.438 | 0.000 - 0.296 | |
| **Mono** | | | 0.053 |
| Mean (SD) | 0.213 (0.083) | 0.252 (0.090) | |
| Range | 0.044 - 0.412 | 0.096 - 0.426 | |
| **B** | | | 0.045 |
| Mean (SD) | 0.148 (0.030) | 0.135 (0.024) | |
| Range | 0.094 - 0.220 | 0.087 - 0.182 | |
| **NK** | | | 0.811 |
| Mean (SD) | 0.108 (0.055) | 0.106 (0.051) | |
| Range | 0.010 - 0.271 | 0.007 - 0.209 | |
| **DC** | | | 0.143 |
| Mean (SD) | 0.016 (0.009) | 0.019 (0.010) | |
| Range | 0.001 - 0.044 | 0.002 - 0.048 | |

*Suppl. Table 3: Imputed cell proportions in PBMCs of late Stable and CAMR patients*

Proportions of cell types in our Stable and CAMR PBMC samples estimated by deconvolution (see Methods). Cell estimations are represented as mean, standard deviation (SD) and range (minimum to maximum value). Comparison between both groups was tested with an ANOVA t-test with equal variances. No cell type exhibited a significant difference between the Stable and CAMR groups after accounting for multiple testing (Bonferroni threshold $p<8.3x10^{-3}$).

*Mono, monocytes; DC, dendritic cells.*

| | Stable (N=43) | ABMR (N=35) | p value |
|---|---|---|---|
| **CD4 TCM** | | | 0.352 |
| Mean (SD) | 0.151 (0.070) | 0.137 (0.063) | |
| Range | 0.000 - 0.316 | 0.035 - 0.302 | |
| **CD8 TCM** | | | 0.094 |
| Mean (SD) | 0.040 (0.038) | 0.026 (0.032) | |
| Range | 0.000 - 0.137 | 0.000 - 0.098 | |
| **CD14 Mono** | | | 0.071 |
| Mean (SD) | 0.172 (0.066) | 0.201 (0.073) | |
| Range | 0.038 - 0.360 | 0.075 - 0.366 | |
| **MAIT** | | | 0.529 |
| Mean (SD) | 0.008 (0.020) | 0.011 (0.023) | |
| Range | 0.000 - 0.096 | 0.000 - 0.083 | |
| **CD4 Naive** | | | 0.980 |
| Mean (SD) | 0.051 (0.025) | 0.051 (0.020) | |
| Range | 0.000 - 0.095 | 0.020 - 0.108 | |
| **ILC** | | | 0.035 |
| Mean (SD) | 0.017 (0.006) | 0.014 (0.007) | |
| Range | 0.000 - 0.027 | 0.003 - 0.024 | |
| **Eryth** | | | 0.363 |
| Mean (SD) | 0.000 (0.001) | 0.001 (0.001) | |
| Range | 0.000 - 0.006 | 0.000 - 0.006 | |
| **CD16 Mono** | | | 0.097 |
| Mean (SD) | 0.037 (0.031) | 0.050 (0.037) | |
| Range | 0.000 - 0.106 | 0.000 - 0.131 | |
| **B naive** | | | 0.017 |
| Mean (SD) | 0.046 (0.017) | 0.037 (0.016) | |
| Range | 0.017 – 0.093 | 0.009 – 0.068 | |
| **CD8 Naïve** | | | 0.963 |
| Mean (SD) | 0.017 (0.028) | 0.017 (0.028) | |
| Range | 0.000 – 0.100 | 0.000 – 0.094 | |
| **Plasmablast** | | | 0.556 |
| Mean (SD) | 0.002 (0.002) | 0.002 (0.003) | |
| Range | 0.000 – 0.008 | 0.000 – 0.011 | |
| **Platelet** | | | 0.512 |
| Mean (SD) | 0.012 (0.009) | 0.013 (0.011) | |

| | Stable (N=43) | ABMR (N=35) | p value |
|---|---|---|---|
| Range | 0.000 – 0.034 | 0.003 – 0.052 | |
| **CD8 TEM** | | | 0.962 |
| Mean (SD) | 0.078 (0.058) | 0.079 (0.050) | |
| Range | 0.000 – 0.268 | 0.000 – 0.181 | |
| **Treg** | | | 0.416 |
| Mean (SD) | 0.031 (0.013) | 0.029 (0.008) | |
| Range | 0.010 – 0.071 | 0.017 – 0.049 | |
| **CD4 Proliferating** | | | 0.122 |
| Mean (SD) | 0.000 (0.001) | 0.002 (0.005) | |
| Range | 0.000 – 0.006 | 0.000 – 0.024 | |
| **NK Proliferating** | | | 0.094 |
| Mean (SD) | 0.000 (0.001) | 0.001 (0.002) | |
| Range | 0.000 – 0.003 | 0.000 – 0.007 | |
| **cDC2** | | | 0.074 |
| Mean (SD) | 0.004 (0.008) | 0.002 (0.004) | |
| Range | 0.000 – 0.030 | 0.000 – 0.018 | |
| **CD4 CTL** | | | 0.524 |
| Mean (SD) | 0.023 (0.023) | 0.026 (0.025) | |
| Range | 0.000 – 0.071 | 0.000 – 0.077 | |
| **HSPC** | | | 0.002 |
| Mean (SD) | 0.005 (0.002) | 0.006 (0.002) | |
| Range | 0.001 - 0.009 | 0.001 - 0.011 | |
| **B intermediate** | | | 0.581 |
| Mean (SD) | 0.048 (0.014) | 0.050 (0.015) | |
| Range | 0.012 - 0.080 | 0.021 - 0.087 | |
| **B memory** | | | 0.966 |
| Mean (SD) | 0.016 (0.012) | 0.016 (0.013) | |
| Range | 0.000 - 0.040 | 0.000 - 0.038 | |
| **CD4 TEM** | | | 0.818 |
| Mean (SD) | 0.007 (0.010) | 0.007 (0.008) | |
| Range | 0.000 - 0.035 | 0.000 - 0.026 | |
| **gdT** | | | 0.266 |
| Mean (SD) | 0.000 (0.001) | 0.001 (0.001) | |
| Range | 0.000 - 0.002 | 0.000 - 0.005 | |
| **NK** | | | 0.241 |
| Mean (SD) | 0.180 (0.049) | 0.166 (0.049) | |
| Range | 0.096 - 0.317 | 0.058 - 0.271 | |

| | Stable (N=43) | ABMR (N=35) | p value |
|---|---|---|---|
| **CD8 Proliferating** | | | 0.701 |
| Mean (SD) | 0.001 (0.000) | 0.001 (0.001) | |
| Range | 0.001 - 0.002 | 0.000 - 0.002 | |
| **dnT** | | | 0.467 |
| Mean (SD) | 0.050 (0.012) | 0.052 (0.011) | |
| Range | 0.026 - 0.081 | 0.028 - 0.077 | |
| **NK_CD56bright** | | | 0.804 |
| Mean (SD) | 0.003 (0.006) | 0.003 (0.005) | |
| Range | 0.000 - 0.027 | 0.000 - 0.024 | |

*Suppl. Table 4: Deep resolution cell type deconvolution of the bulk RNA-seq data for late Stable and CAMR patients*

Cell estimates are presented as mean, standard deviation (SD) and range (minimum to maximum value). Continuous values were tested with an ANOVA (t-test with equal variances). No cell type was significantly different between both groups when accounting for multiple testing (Bonferroni threshold $p=1.9x10^{-3}$).

*Mono, monocyte; MAIT, mucosal associated invariant T cell; ILC, Innate lymphoid cell; Eryth, erythrocyte; TEM, T effector memory cell; cDC2, classical dendritic cell 2; CTL, cytotoxic T lymphocyte; HSPC, hematopoietic stem and progenitor cell; gdT, gamma-delta T cell; dnT, double negative T cell.*

| Gene | Stat | Pval | FDR | AveFC | propDataset |
|---|---|---|---|---|---|
| **E2F5** | 4.66 | 1.60E-06 | 0.0002 | 0.08 | 0.83 |
| **HBB** | 4.56 | 2.57E-06 | 0.0002 | -0.10 | 1.00 |
| **TLN1** | 4.42 | 4.95E-06 | 0.0003 | 0.19 | 0.83 |
| **MLH3** | 3.96 | 3.76E-05 | 0.0015 | 0.02 | 0.83 |
| **C1QB** | 3.94 | 4.11E-05 | 0.0015 | 0.72 | 0.83 |
| **PLCB1** | 3.63 | 0.0001 | 0.0045 | -0.15 | 1.00 |
| **SH3PXD2B** | 3.46 | 0.0003 | 0.0073 | 0.07 | 0.83 |
| **LST1** | 3.42 | 0.0003 | 0.0073 | 0.17 | 0.83 |
| **PSMD9** | 3.23 | 0.0006 | 0.0128 | 0.08 | 0.83 |
| **TYROBP** | 3.00 | 0.0013 | 0.0221 | 0.00 | 1.00 |
| **ERG** | 3.00 | 0.0014 | 0.0221 | 0.17 | 1.00 |
| **C1QA** | 2.97 | 0.0015 | 0.0221 | 0.44 | 0.83 |
| **CEACAM8** | 2.96 | 0.0015 | 0.0221 | 0.51 | 0.83 |
| **PSMB6** | 2.91 | 0.0018 | 0.0226 | 0.15 | 0.83 |
| **NFAM1** | 2.91 | 0.0018 | 0.0226 | 0.04 | 0.83 |
| **CFL1** | 2.85 | 0.0022 | 0.0240 | 0.00 | 0.83 |
| **MMP9** | 2.83 | 0.0023 | 0.0240 | 0.04 | 1.00 |
| **CHIT1** | 2.81 | 0.0025 | 0.0242 | 0.31 | 0.83 |
| **STS** | 2.79 | 0.0026 | 0.0242 | 0.10 | 1.00 |
| **VASP** | 2.78 | 0.0028 | 0.0242 | 0.04 | 0.83 |
| **CEBPE** | 2.77 | 0.0028 | 0.0242 | 0.10 | 0.83 |
| **PGLYRP1** | 2.74 | 0.0031 | 0.0247 | 0.27 | 0.67 |
| **ADAMTS2** | 2.73 | 0.0032 | 0.0247 | 0.12 | 0.83 |
| **MVP** | 2.72 | 0.0033 | 0.0248 | 0.09 | 0.67 |
| **MS4A3** | 2.60 | 0.0046 | 0.0334 | 0.23 | 0.67 |
| **GNPDA2** | 2.56 | 0.0053 | 0.0351 | 0.00 | 0.67 |
| **CHI3L1** | 2.55 | 0.0053 | 0.0351 | 0.31 | 1.00 |
| **IL24** | 2.55 | 0.0054 | 0.0351 | 0.08 | 1.00 |
| **BEND4** | 2.52 | 0.0058 | 0.0365 | 0.23 | 0.67 |
| **C1QC** | 2.48 | 0.0065 | 0.0385 | 0.72 | 0.83 |
| **PLEKHA1** | 2.45 | 0.0071 | 0.0405 | -0.22 | 0.83 |
| **PRKACA** | 2.44 | 0.0074 | 0.0410 | 0.10 | 0.83 |
| **INHBA** | 2.37 | 0.0090 | 0.0482 | 0.33 | 1.00 |
| **CRISP3** | 2.34 | 0.0096 | 0.0502 | 0.30 | 0.67 |
| **MGAM** | 2.32 | 0.0101 | 0.0511 | 0.03 | 1.00 |
| **NUP107** | 2.30 | 0.0108 | 0.0533 | 0.05 | 0.67 |
| **FLT3** | 2.21 | 0.0135 | 0.0644 | -0.01 | 1.00 |
| **ZYX** | 2.20 | 0.0139 | 0.0644 | 0.17 | 0.83 |
| **PTPRJ** | 2.19 | 0.0142 | 0.0644 | 0.01 | 1.00 |
| **XPO4** | 2.18 | 0.0145 | 0.0644 | 0.18 | 0.67 |
| **CEACAM6** | 2.16 | 0.0155 | 0.0663 | 0.28 | 0.83 |
| **FGF13** | 2.15 | 0.0159 | 0.0663 | 0.00 | 0.83 |
| **DAP** | 2.14 | 0.0163 | 0.0665 | 0.01 | 1.00 |
| **ZFP82** | 2.12 | 0.0170 | 0.0680 | 0.05 | 0.67 |
| **LILRB3** | 2.09 | 0.0183 | 0.0715 | -0.03 | 0.83 |

| | | | | | |
|---|---|---|---|---|---|
| **WAS** | 2.02 | 0.0219 | 0.0839 | 0.20 | 0.67 |
| **CLIC1** | 1.99 | 0.0231 | 0.0867 | 0.09 | 0.83 |
| **HK1** | 1.95 | 0.0256 | 0.0942 | -0.01 | 0.83 |
| **TKT** | 1.94 | 0.0265 | 0.0942 | -0.03 | 1.00 |
| **SAP30** | 1.93 | 0.0267 | 0.0942 | -0.25 | 0.83 |
| **FAM111B** | 1.92 | 0.0274 | 0.0942 | -0.09 | 0.67 |
| **PNPLA2** | 1.92 | 0.0276 | 0.0942 | 0.08 | 0.67 |
| **INSR** | 1.87 | 0.0307 | 0.1031 | 0.07 | 1.00 |
| **SPTB** | 1.83 | 0.0336 | 0.1103 | 0.13 | 0.83 |
| **ERGIC1** | 1.82 | 0.0342 | 0.1103 | 0.05 | 0.67 |
| **ABCA13** | 1.82 | 0.0346 | 0.1103 | 0.26 | 0.67 |
| **SH3YL1** | 1.80 | 0.0361 | 0.1130 | 0.01 | 0.67 |
| **SLAMF1** | 1.77 | 0.0380 | 0.1156 | -0.06 | 1.00 |
| **ZNF273** | 1.77 | 0.0385 | 0.1156 | -0.06 | 0.67 |
| **NUDT16** | 1.77 | 0.0387 | 0.1156 | 0.07 | 0.83 |
| **RPRD2** | 1.73 | 0.0419 | 0.1213 | 0.08 | 0.67 |
| **RILPL2** | 1.71 | 0.0435 | 0.1239 | 0.00 | 0.67 |

*Suppl. Table 5: Summary statistics for the meta-analysis in six independent cohorts assessing differential expression in Stable (n=360) vs. CR (n=131) patients*

Only the 188 DEG identified in the discovery cohort were tested in this meta-analysis. Stat corresponds to the statistics calculated by the Stouffer method, AveFC is the average fold change between the six datasets, and the propDataset corresponds to the proportion of the datasets in which data for the tested gene were available.

| Gene | baseMean | log2FoldChange | lfcSE | stat | pvalue | padj |
|---|---|---|---|---|---|---|
| **MMP9** | 1059.37 | -3.38 | 0.64 | -5.27 | 1.35E-07 | 0.0013 |
| **CNTNAP3** | 416.92 | -4.17 | 0.81 | -5.14 | 2.70E-07 | 0.0013 |
| **CXCL1** | 570.64 | -3.66 | 0.71 | -5.13 | 2.89E-07 | 0.0013 |
| **MME** | 2434.75 | -3.12 | 0.62 | -5.04 | 4.67E-07 | 0.0015 |
| **MGAM2** | 164.51 | -3.42 | 0.73 | -4.71 | 2.43E-06 | 0.0062 |
| **BMX** | 153.36 | -3.49 | 0.75 | -4.68 | 2.84E-06 | 0.0062 |
| **IL27** | 98.28 | -1.30 | 0.28 | -4.63 | 3.73E-06 | 0.0070 |
| **CXCR1** | 1381.07 | -2.78 | 0.62 | -4.52 | 6.32E-06 | 0.0094 |
| **PLIN5** | 114.94 | -2.38 | 0.53 | -4.49 | 7.03E-06 | 0.0094 |
| **MGAM** | 4236.05 | -2.45 | 0.55 | -4.48 | 7.58E-06 | 0.0094 |
| **METTL5** | 171.02 | 0.54 | 0.12 | 4.43 | 9.52E-06 | 0.0094 |
| **IL1R2** | 1997.55 | -3.00 | 0.68 | -4.39 | 1.13E-05 | 0.0094 |
| **CYP4F3** | 1843.36 | -2.49 | 0.57 | -4.39 | 1.14E-05 | 0.0094 |
| **PPP1R9A** | 111.47 | 1.40 | 0.32 | 4.39 | 1.14E-05 | 0.0094 |
| **CXCR2** | 3085.73 | -2.57 | 0.59 | -4.38 | 1.18E-05 | 0.0094 |
| **ADGRG3** | 1526.89 | -2.37 | 0.54 | -4.38 | 1.19E-05 | 0.0094 |
| **IL12RB2** | 392.98 | 0.87 | 0.20 | 4.38 | 1.21E-05 | 0.0094 |
| **FFAR2** | 4053.54 | -2.18 | 0.50 | -4.35 | 1.39E-05 | 0.0101 |
| **FCGR3B** | 7538.70 | -3.03 | 0.70 | -4.34 | 1.46E-05 | 0.0101 |
| **MMP25** | 2718.69 | -1.87 | 0.44 | -4.26 | 2.09E-05 | 0.0138 |
| **BTBD19** | 205.94 | -1.21 | 0.29 | -4.21 | 2.53E-05 | 0.0159 |
| **LOC400499** | 862.45 | -2.25 | 0.54 | -4.20 | 2.68E-05 | 0.0161 |
| **TUFT1** | 73.49 | -1.07 | 0.26 | -4.17 | 3.01E-05 | 0.0166 |
| **NAMPT** | 32458.89 | -1.53 | 0.37 | -4.16 | 3.12E-05 | 0.0166 |
| **KCNJ15** | 1604.47 | -2.35 | 0.57 | -4.16 | 3.14E-05 | 0.0166 |
| **TNFRSF10C** | 1887.00 | -2.21 | 0.53 | -4.15 | 3.34E-05 | 0.0170 |
| **DAAM2** | 174.77 | -3.42 | 0.84 | -4.08 | 4.53E-05 | 0.0222 |
| **PTGS2** | 6000.52 | -2.05 | 0.50 | -4.06 | 4.90E-05 | 0.0230 |
| **AATK** | 429.39 | -1.40 | 0.34 | -4.05 | 5.05E-05 | 0.0230 |
| **PANX2** | 141.49 | -1.57 | 0.39 | -4.04 | 5.27E-05 | 0.0232 |
| **KY** | 713.67 | -2.70 | 0.67 | -4.03 | 5.62E-05 | 0.0240 |
| **PGLYRP1** | 301.26 | -2.42 | 0.60 | -4.01 | 6.14E-05 | 0.0248 |
| **TREM1** | 11853.13 | -1.43 | 0.36 | -4.01 | 6.20E-05 | 0.0248 |
| **LONRF3** | 651.14 | -0.69 | 0.17 | -3.97 | 7.18E-05 | 0.0279 |
| **DCST1** | 32.35 | -1.52 | 0.38 | -3.95 | 7.78E-05 | 0.0294 |
| **LRG1** | 281.46 | -1.64 | 0.42 | -3.94 | 8.17E-05 | 0.0300 |
| **PROK2** | 2025.03 | -1.98 | 0.51 | -3.91 | 9.21E-05 | 0.0329 |
| **GNG10** | 61.75 | -1.31 | 0.34 | -3.88 | 0.0001 | 0.0364 |
| **CXCL8** | 19293.56 | -2.73 | 0.72 | -3.81 | 0.0001 | 0.0472 |
| **PRDM5** | 120.18 | -1.95 | 0.52 | -3.78 | 0.0002 | 0.0499 |
| **TRIM9** | 26.41 | -2.05 | 0.54 | -3.78 | 0.0002 | 0.0499 |
| **CACNA1E** | 131.99 | -2.53 | 0.67 | -3.77 | 0.0002 | 0.0499 |
| **RHBDL1** | 21.04 | -1.42 | 0.38 | -3.77 | 0.0002 | 0.0499 |
| **NCF4** | 1678.95 | -0.91 | 0.24 | -3.76 | 0.0002 | 0.0499 |
| **G0S2** | 3486.31 | -2.76 | 0.74 | -3.75 | 0.0002 | 0.0499 |

| | | | | | | |
|---|---|---|---|---|---|---|
| **MANSC1** | 144.64 | -2.03 | 0.54 | -3.75 | 0.0002 | 0.0499 |

*Suppl. Table 6: Summary statistics for the DEG between 1-year stable reference patients who remained stable (n=51) and those who later experienced a post-1-year CAMR event (n=11)*

baseMean, base mean expression; log2FoldChange, log2 transformation of the fold change between CAMR and Stable patients; lfcSE, log2(Fold change) standard error; stat, Wald statistic.
